# Explicit knowledge of task structure is the primary determinant of human model-based action

**DOI:** 10.1101/2020.09.06.20189241

**Authors:** Pedro Castro-Rodrigues, Thomas Akam, Ivar Snorasson, M Marta Camacho, Vitor Paixão, J. Bernardo Barahona-Corrêa, Peter Dayan, H. Blair Simpson, Rui M. Costa, Albino J. Oliveira-Maia

**Author notes:** Equally contributing authors (Pedro Castro-Rodrigues, Thomas Akam). John Van Geest Center for Brain Repair, University of Cambridge, UK.

## Abstract

Explicit information obtained through instruction profoundly shapes human choice behaviour. However, this has been studied in computationally simple tasks, and it is unknown how model-based and model-free systems, respectively generating goal-directed and habitual actions, are affected by the absence or presence of instructions. We assessed behaviour in a novel variant of a computationally more complex decision-making task, before and after providing information about task structure, both in healthy volunteers and individuals suffering from obsessive-compulsive (OCD) or other disorders. Initial behaviour was model-free, with rewards directly reinforcing preceding actions. Model-based control, employing predictions of states resulting from each action, emerged with experience in a minority of subjects, and less in OCD. Providing task structure information strongly increased model-based control, similarly across all groups. Thus, explicit task structural knowledge determines human use of model-based reinforcement learning, and is most readily acquired from instruction rather than experience.

## Introduction

The brain uses multiple systems to choose which actions to perform^1–6^. One widely held distinction is made between goal-directed actions, guided by predictions of their specific outcomes, and habitual actions, performed according to preferences acquired through prior reinforcement^1,7–9^. This cognitive and behavioural classification is thought to correspond, at least in part, to a computational distinction between two different types of reinforcement learning (RL), termed model-based and model-free^4,5,10,11^. Model-based RL learns to predict the specific consequences of actions, and computes their values, i.e. long run utilities, by simulating likely future behavioural trajectories. This allows for statistically efficient use of experience, and thus behavioural flexibility, at the cost of the computational demands of planning. Model-free RL, by contrast, learns estimates of the value of states or actions directly from experience, and updates these estimates using reward prediction errors. This allows for rapid action selection at low computational cost, but uses information less efficiently, resulting in slower adaption to changes in the environment. It is thought that the brain takes advantage of the complementary strengths of both prospective (model-based) and retrospective (model-free) approaches to decision-making, through mechanisms that estimate whether the payoff for more accurate prediction is worth the computational costs of planning^4,12,13^.

Sequential, or multi-step, decision tasks have emerged as a powerful approach to study model-based and model-free RL in humans^11,12,14,15^. In such tasks, subjects move through a sequence of states to obtain rewards, typically with non-stationary reward and/or action-state transition probabilities, forcing continuous learning. The contributions of model-based and model-free RL can be determined by examining how subjects update their choices in light of recent experience. To date, the most commonly used task is the ‘two-step’ task, employing a choice between two ‘first-step’ stimuli, which leads probabilistically to one of two ‘second-step’ states, where rewards may be obtained^11^. Each first-step stimulus commonly leads to one of the second-step states but, on a minority of trials, leads to the state commonly reached from the other stimulus. Model-based and model-free RL are identified according to how the trial outcome (rewarded or not) and state transition (common or rare) interact to affect the subsequent choice. Under model-free control, the agent will tend to repeat first-step choices that are followed by reward, irrespective of the state transition. By contrast, under model-based control, the agent will tend to switch first-step choice when a rare transition leads to reward, as the reward increases the value of the state commonly reached from the not-chosen first step option. The two-step task has been used to study neural correlates of model-based and model-free control in healthy subjects^11,16,25,26,17–24^, and to investigate decision making in clinical populations^27–31^.

Human subjects typically receive extensive instruction about task structure prior to performing the two-step task^11,32^. However, and even though there is extensive literature showing that instruction profoundly shapes human behaviour in operant^33–35^ and fear^36,37^ conditioning, as well as value-based decision making^38–41^, little is known about how instruction affects behaviour in multi-step tasks that dissociate model-based and model-free control. To our knowledge, a single study in healthy humans has partially addressed this question, showing that making instructions more comprehensive and easier to understand, increases the influence of model-based relative to model-free RL^32^. This result, in combination with other analyses, led the authors to propose that humans are primarily model-based learners on this task, suggesting that apparent model-free behaviour, including in some clinical populations, may result from incorrect task models, rather than a true model-free strategy.

However, no studies have explored behaviour on multi-step tasks in the absence of information about task structure, in either healthy or clinical populations. Thus, it remains unclear how model-based and model-free RL contribute to action selection in situations where subjects must learn task structure directly and exclusively from experience, and how providing explicit information about task structure affects each system. To address these questions, we created a new version of the two-step task, requiring minimal prior instruction and we initially administered it with no information given about the task state space, transition structure, or reward probabilities. Then, the task was repeated following debriefing about these elements of the task’s structure. Behaviour was tested in healthy volunteers, as well as in a sample of individuals with obsessive-compulsive disorder (OCD), previously reported to have deficits in the degree to which model-based RL is employed^28,30^. To control for the effects of psychotropic medication and of unspecific mood and anxiety symptoms, data was also collected in a comparison sample of individuals with mood and anxiety disorders. Initial behaviour, prior to instructions, was model-free across all groups, with model-based control emerging with experience in only a minority of subjects, and to a lesser extent in OCD. However, once task structure information was provided, model-based control increased to a very similar, and significant extent in healthy volunteers and individuals with OCD or other disorders. These findings support the conclusion that explicit task structural knowledge determines human use of model-based RL, and is most readily acquired from instruction rather than experience.

## Results

We developed a two-step task requiring minimal prior instruction, which we name ‘simplified two-step task’, by simplifying the visual representation of task states on the screen, the task structure (allowing only a single action rather than a choice in each second-step state), and the reward probability distribution (using blocks, instead of slowly fluctuating Gaussian random walks, to increase the contrast between good and bad options^42,43^; Figure 1). Two-hundred and four individuals were recruited in Lisbon and New York to perform this task: 109 were healthy volunteers, 46 were diagnosed with OCD and 49 with other mood and anxiety disorders. Sociodemographic and psychometric data from all participants is shown in table 1. While differences between groups were not found for age (F=2.4, *P*=0.1; one-way ANOVA) nor gender (χ^2^=5.1, *P*=0.3; Pearson’s chi-squared), they were present for years of education (F=5.7, *P*<0.01; one-way ANOVA), which were only slightly, but significantly, higher in healthy volunteers than the mood and anxiety group (*P*=0.01; Tukey’s HSD). As expected, both clinical groups had significantly higher anxiety and depression scores than healthy volunteers (F>55, *P*<0.001; across all one-way ANOVA’s for the depression and anxiety scales), while participants with OCD had higher obsessive-compulsive scores than participants in either of the two other groups (F>200; *P*<0.001; across all one-way ANOVA’s for the Yale-Brown Obsessive-Compulsive Scale total and sub-scores).

**Figure 1.**
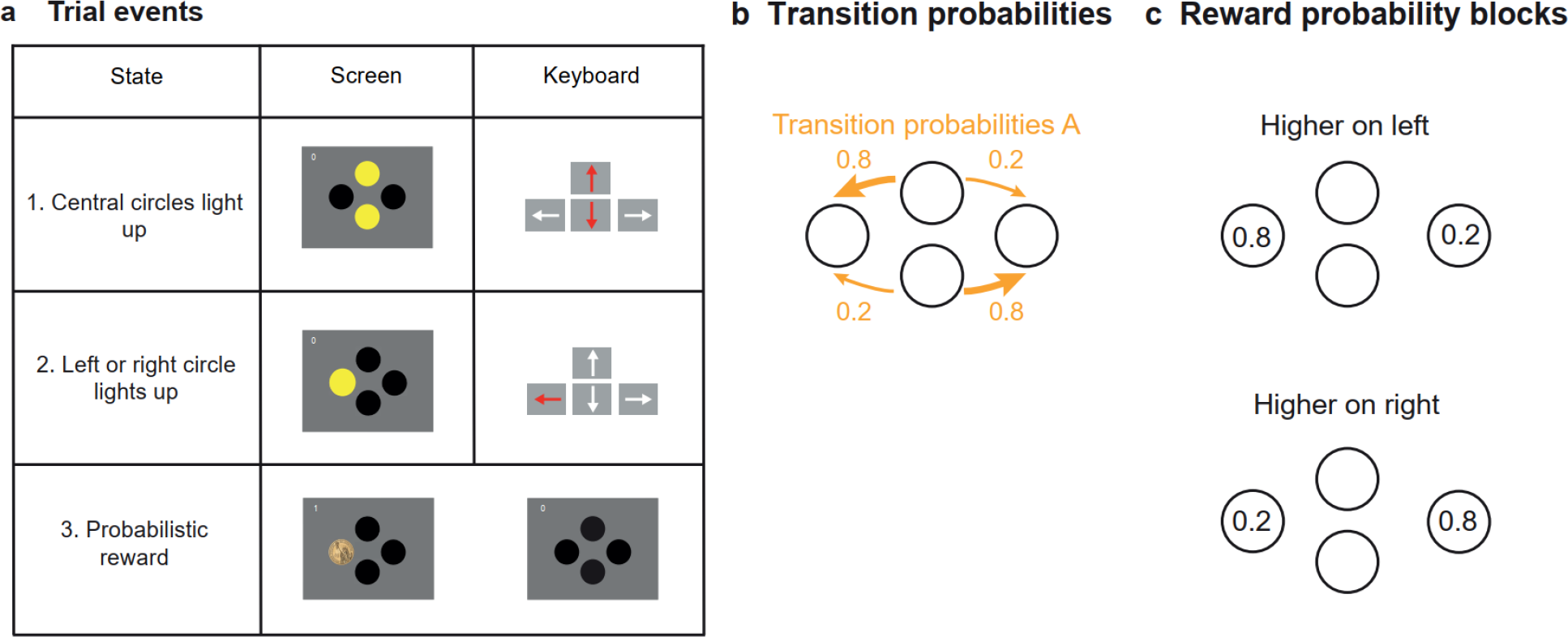
Behavioural Task. The simplified two-step task was presented on a computer screen with 4 circles visible on a grey background: 2 central circles (upper and lower) and two side circles (right and left). Each circle was coloured yellow when available for selection, and black when unavailable. Circles could be selected by pressing the corresponding arrow key (up, down, left or right) on the computer keyboard. **a)** Trial events. Each trial started with the central circles turning yellow, prompting the choice between either upper or lower circle (a1). Following the first-step choice, one of the side circles (left or right) would become yellow (a2), with differing probabilities (also see b). The subject would then select the yellow side circle resulting in a probabilistic monetary reward. Reward was indicated by the side circle changing to the image of a coin (a3 left) while no reward was indicated by the circle changing back to black (a3 right; also see c). **b)** Transition probabilities. At each trial, choosing one circle at the first step commonly (p=0.8) lit up one side circle and rarely (p=0.2) the other side circle, with inverse probabilities for choosing the alternate circle. Type A transition probabilities (common transitions from upper to left and from lower to right) are shown here. Reverse transition probabilities (Type B) are the alternate possibility. The transition probabilities were fixed (either A or B, counterbalanced across subjects) in the Fixed version of the task, or underwent reversals from A to B in the Changing version (see methods and supplementary results for additional details on this version). **c)** Reward probability blocks. The reward probabilities upon selection of the side circles changed in blocks that were either neutral (p=0.4 for both left and right sides), or higher on one of the sides (p=0.8 vs p=0.2, i.e., non-neutral blocks), as shown here. Non-neutral blocks ended when subjects consistently chose the first-step option (upper or lower) that most frequently lead to the high reward probability side. Neutral blocks ended probabilistically, independent of subjects’ behaviour (see methods for details). To maximize reward rate, subjects must choose the first step action which commonly leads to the second-step state with higher reward probability, tracking the best option across reward-probability reversals.

**Table 1.** Sociodemographic and psychometric characterization of study samples.

|  | **HV** | **OCD** | **MA** |
| --- | --- | --- | --- |
| Sex (% males) | 33% | 41% | 31% |
| Age (years) | 30.4 (7.1) | 34.1 (12.4) | 32.6 (13.1) |
| Education (years completed) | 16.2 (2.5) | 15.1 (2.9) | 14.5 (4.1) |
| YBOCS total score | 1.5 (3.5) | 23 (6.4) | 2.7 (4.8) |
| Y-BOCS obsessions | 0.6 (1.8) | 11 (3.3) | 2.1 (3.7) |
| Y-BOCS compulsions | 0.9 (1.9) | 12 (3.5) | 0.6 (1.9) |
| STAI-state score | 31.5 (8.1) | 47.6 (15.4) | 47.9 (11.3) |
| STAI-trait score | 30.8 (8) | 56.6 (12) | 53.1 (10.1) |
| BDI-II score<sup>a</sup> | 4 (4.8) | 21.1 (16.2) | 24.8 (12.1) |
| DASS depression score<sup>b</sup> | 1.5 (1.8) | 7.8 (5.6) | 7.9 (4.4) |
| DASS anxiety score<sup>b</sup> | 0.6 (1.2) | 5.2 (4.4) | 5.8 (4.3) |
| DASS stress score<sup>b</sup> | 2.5 (2.2) | 10.4 (4.7) | 8.5 (4.5) |
| Corsi block tapping test - total span<sup>a</sup> | 16 (3.1) | 15.4 (3.9) | 13.1 (2.5) |
| No-Go errors in Go/No-Go task (n) | 11.2 (7.4) | 16 (12.5) | 21.2 (11.6) |
| Reaction time in Go/No-Go task (ms) | 470 (44.4) | 517 (55.1) | 510 (55.1) |

HV = Healthy volunteers; OCD = Obsessive-compulsive disorder; MA = Mood and anxiety disorders. <sup>a</sup> - only in Lisbon groups; <sup>b</sup> - only in New York groups; YBOCS-II = Yale-Brown Obsessive-Compulsive Scale-II; STAI = State-Trait Anxiety Inventory; BDI-II = Beck Depression Inventory; DASS = Depression Anxiety Stress Scales. Data presented are mean values (standard deviations).

While developing the task among healthy volunteers in Lisbon, participants were randomized between two different versions, one with fixed transition probabilities linking the first-step actions and second-step states (Fixed version; n=40), and one where the transition probabilities underwent periodic reversals (Changing version; n=42). The Changing version proved too complex for most subjects (see supplementary results for details) so we subsequently focused on the Fixed task, which is used for all data and figures in the main text. All healthy controls recruited in New York (n=27), and all clinical subjects at both sites (n=95) were run on this version. All subjects in both versions performed 4 sessions of 300 trials each in a single day. A subset of healthy controls, and all clinical subjects, were debriefed between sessions 3 and 4, with the task structure explained to them. We assessed the effect of uninstructed experience by comparing behaviour between sessions 1 and 3, and the effect of explicit knowledge by comparing behaviour between sessions 3 and 4.

### Initial behaviour is under model-free control

As subjects were not told how their actions (arrow key presses) affected the stimuli shown on the screen, they had to learn both the correspondence between arrow keys and stimuli, and that stimuli could only be selected when highlighted. In the 67 healthy volunteers performing the Fixed version of the task, the number of invalid key presses per trial (i.e. presses to keys whose corresponding stimulus was not highlighted) decreased over the first 50-100 trials, before stabilising at a low level in all but a minority of subjects (Figure S1). To assess how trial events affected subject’s choices, we analysed the probability of repeating first-step choices (termed ‘stay probabilities’) as a function of the previous state transition (common or rare), trial outcome (rewarded or not), and their interaction^11^. During session 1, stay probability was strongly influenced by trial outcome (*P*<0.0001, permutation test), but not by state transition (*P*=0.3) nor the transition-outcome interaction (*P*=0.1; Figure 2 a, d). This pattern is consistent with a simple model-free strategy, in which the outcome received at the end of the trial directly reinforces the choice made at the first-step^11^. This direct reinforcing effect of reward was evident from the very start of the first session (Figure 2b), rather than emerging with task experience. Although state transitions in session 1 did not influence subsequent first-step choices, key-press reaction times at the second-step were faster following common than rare transitions (399.1 ± 16.9ms and 514.4 ± 20.5ms respectively; t_66_=7.81, *P*<0.0001, paired t-test; Figure 2e). This dissociation may reflect motor systems learning to predict and prepare upcoming actions before decision making systems are using a predictive model to evaluate choices.

**Figure 2.**
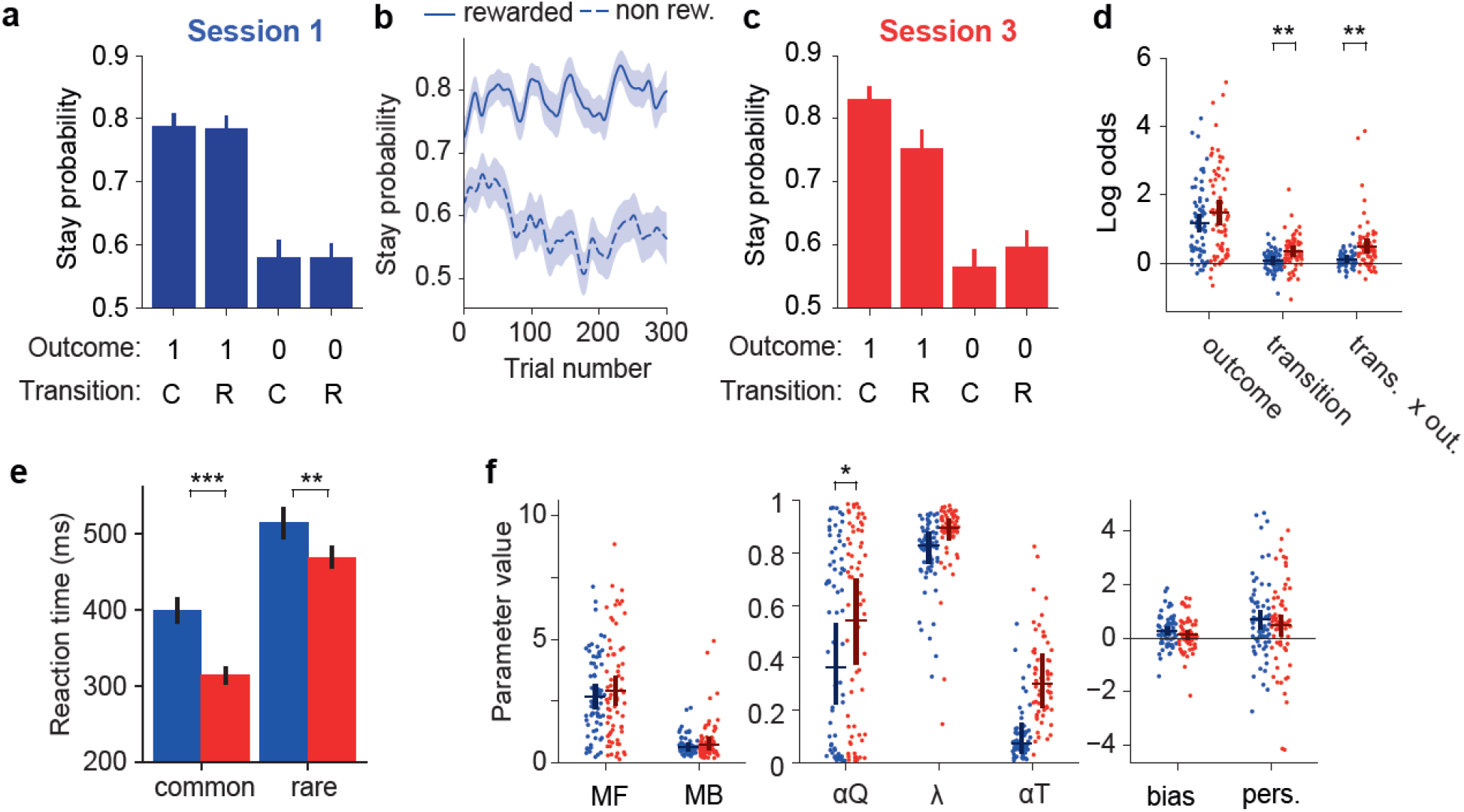
Uninstructed behaviour is predominantly model-free. **a)** Session 1 stay probability analysis showing the probability of repeating the first step choice on the next trial as a function of trial outcome (rewarded or not rewarded) and state transition (common or rare) across 67 healthy volunteers. Error bars indicate across subject standard error (SEM). **b)** Stay probability for rewarded and non-rewarded trials as a function of trial number in session 1. Shaded area shows across subject standard error. **c)** Stay probabilities for session 3. **d)** Logistic regression analysis of how the outcome (rewarded or not), transition (common or rare) and their interaction, predict the probability of repeating the same choice on the subsequent trial. Dots indicate maximum a posteriori parameter values for individual subjects, bars indicate the population mean and 95% confidence interval of the mean. In this and other panels, blue indicates session 1 while red indicates session 3. **e)** Reaction times after common and rare transitions in session 1 and 3. **f)** Comparison of mixture model fits between session 1 and session 3. Dots and bars are represented as in panel C. RL model parameters: MF: Model-free strength, MB: Model-based strength, αQ: Value learning rate, λ: Eligibility trace, αT: Transition prob. learning rate, bias: Choice bias, pers.: Choice perseveration. In all figures significant differences are indicated as: * *P*<0.05, ** *P*<0.01, *** *P*<0.001.

### Uninstructed experience has limited effects on model-based control

To assess how task experience affected behavioural strategy we compared behaviour in sessions 1 and 3. Stay probability at session 3 was more strongly influenced by both state transition (*P*=0.004, permutation test) and the transition-outcome interaction (*P*=0.002), while the influence of trial outcome increased only at trend level (*P*=0.06; Figure 2 c, d). This pattern is consistent with increased influence of model-based control, as model-based agents know that outcomes following rare transitions primarily influence the value of the first-step option that was not chosen^11,42^, leading to loading on the transition-outcome interaction predictor. Importantly, loading on the transition–outcome interaction parameter across sessions 1 to 3 was positively correlated with the number of rewards obtained by each subject (rho=0.67, *P*<0.001), suggesting that subjects who learned a model of the task obtained rewards at a higher rate. Furthermore, we have previously shown that, when transition probability estimates are updated based on experienced state transitions, as is the case here, model-based agents tend to repeat the same choice after common transitions, producing a positive coefficient for state transition as a predictor of stay probability^42^. Increased loading on the state transition predictor thus provides added support for development of model-based control with task experience between sessions 1 and 3. While key-press reaction times at the second-step became faster overall between session 1 and 3 (main effect of session *P*<0.0001, repeated measures ANOVA), this was more pronounced following common than rare transitions (session-transition interaction *P*=0.008; Figure 2e).

To further explore model-free and model-based control prior to receiving instructions, we fit RL models to data from sessions 1-3. Model-comparison combining data from sessions 1-3 indicated that a mixture model including model-free and model-based components fit data better than a purely model-free or a purely model-based model, as reflected by lower Bayesian Information Criteria (BIC) scores for the mixture model (Figure S2, left panel). Models that included a “bias” parameter, capturing bias towards the upper or lower first-step choice, and a “perseveration” parameter, capturing a tendency to repeat the previous choice, fit the data better than a model not including these parameters (Figure S2, right panel). To assess uninstructed learning effects, we compared parameter values of the fitted models for sessions 1 and 3. The value learning rate increased significantly between session 1 and 3 (*P=*0.04, permutation test), but no other parameter changed significantly, including the strength of model-based control (Figure 2f). The discrepancy with increased loading on the ‘transition x outcome’ predictor in the stay-probability analysis may reflect lower statistical power to detect subtle strategy changes in the strongly non-linear and higher parameter count RL model. This is supported by findings from model comparison for individual subjects between the mixture RL model and a simpler model-free RL model, indicating that only about 15% of subjects (10/67) had learned to use model-based RL at session 3 (likelihood ratio test, threshold *P*=0.05). Overall, this data shows that model-free RL dominated initial human behaviour in this unfamiliar domain and remained the strongest influence on choice behaviour for most subjects over the 900 uninstructed trials.

### In OCD uninstructed behaviour is biased towards model-free control

Ninety-five individuals with either OCD or other mood and anxiety disorders (Table 1) also completed the Fixed version of the simplified two-step task. In the stay probability analysis, when comparing session 1 with session 3, the OCD group (n=46) did not show the increased influence of transition or transition-outcome interaction that was seen in healthy controls (*P*=0.08 and *P*=0.71 respectively; Figure 3a, b). Instead there was an increased influence of trial outcome over uninstructed learning (*P*=0.001), not seen in controls, that may reflect enhanced model-free control with experience. However, in direct comparisons with healthy volunteers (n=67), session by group interaction was significant only for the transition-outcome interaction parameter (*P*=0.01), but not for the transition (P=0.6) or outcome parameters (*P*=0.2).

**Figure 3.**
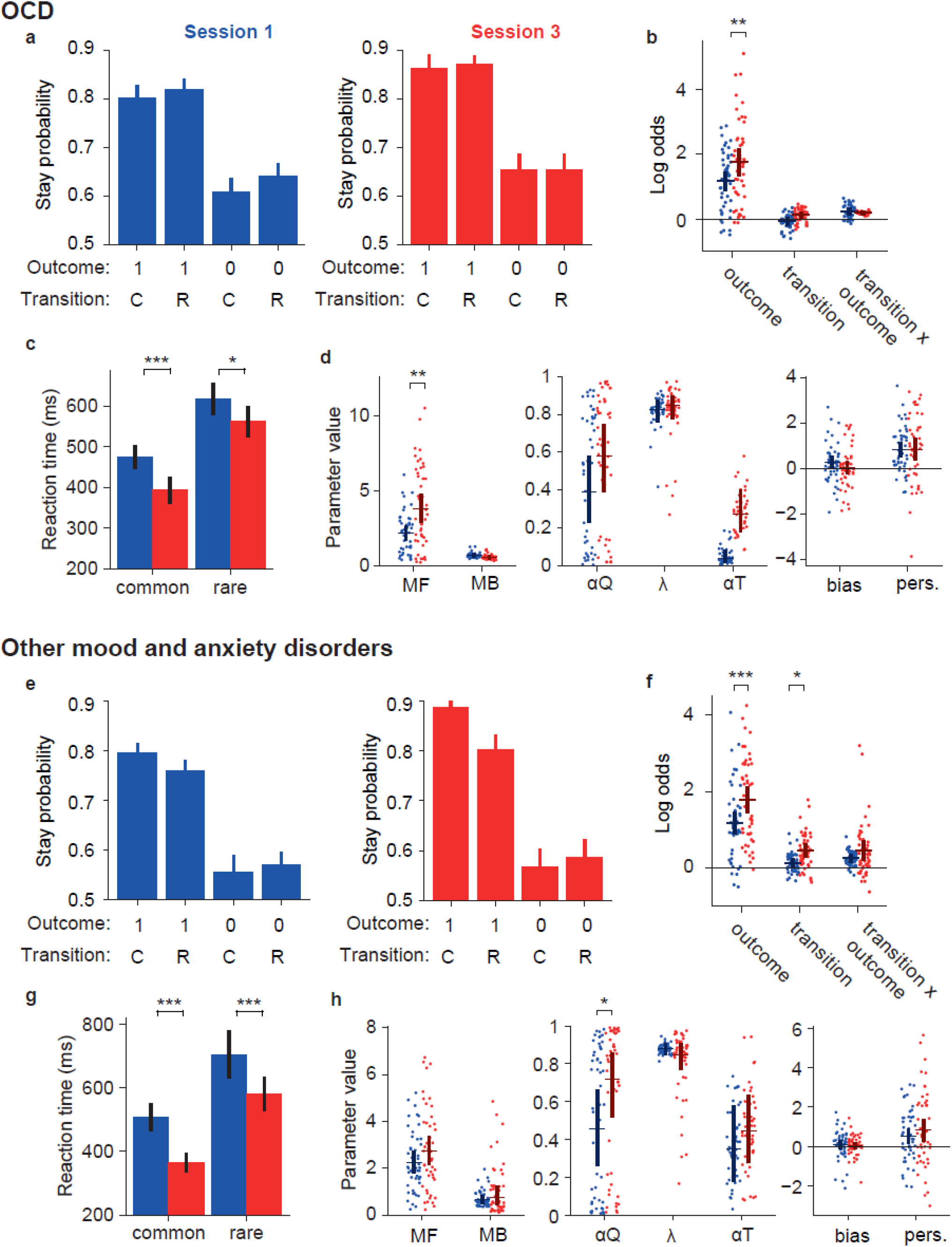
Uninstructed behaviour is biased towards model-free control in OCD. Data for individuals with OCD (n=46) is represented in panels a-c while that for individuals with other mood and anxiety disorders (n=49) are in panels d-f. **(a, d)** Stay probability analysis showing the probability of repeating the first step choice on the next trial as a function of trial outcome (rewarded or not rewarded) and state transition (common or rare). Error bars indicate the cross subject standard error of the mean (SEM). The left (blue) panel shows data from the first session, the right (red) panel shows data from session 3. **(b, e)** Logistic regression analysis of how the outcome (rewarded or not), transition (common or rare) and their interaction, predict the probability of repeating the same choice on the subsequent trial. Dots indicate maximum *a posteriori* loading for individual subjects, bars indicate the population mean and 95% confidence interval on the mean. **(c, f)** Comparison of mixture model fits. Dots and bars are represented as in panels C and G. RL model parameters: MF, Model-free strength; MB, Model-based strength; αQ, Value learning rate; λ, Eligibility trace; αT, Transition probability learning rate; bias, Choice bias; pers., Choice perseveration.

As in healthy subjects, second-step reaction times were faster following common than rare transitions (main effect of transition, *P*<0.0001, repeated measures ANOVA; Figure 3c), and also faster in session 3 than session 1 (main effect of session, *P*=0.003). However, the session by transition interaction did not reach significance (*P*=0.2), indicating that for this group the effect of experience on RT did not differentiate between common and rare transitions. Directly comparing OCD and healthy volunteers, while individuals with OCD had slower reaction times overall (*P*=0.03, linear mixed model), interactions with group were not significant for session (*P*=0.9), transition (*P*=0.4), nor session by transition interaction (*P*=0.7). Finally, consistent with the stay probability analysis, RL mixture model fits to sessions 1 and 3 (Figure 3c) showed an increase in the influence of model-free action values on choice over learning (*P*=0.008, permutation test), that was not seen in healthy volunteers, with a trend toward significant session-by-group interaction (*P*=0.07).

To investigate potential contributions of medication or of unspecific mood and anxiety symptoms for the findings in the OCD group, equivalent experiments and comparisons were performed in a sample of individuals with other mood and anxiety disorders (n=49). Here, in stay probability analysis (Figure 3e, f) we found an increased influence of trial outcome (*P*<0.001) and transition (*P*=0.01) with task experience, but not the transition-outcome interaction predictor (*P*=0.7). Second-step reaction times were faster following common than rare transitions (main effect of transition, *P*<0.0001, Figure 3g), and faster in session 3 than session 1 (main effect of session, *P*<0.0001), but the session by transition interaction was not significant (*P*=0.4). Compared with the healthy volunteers, this group had slower second-step reaction times overall (main effect of group, *P*=0.03), but particularly on rare transition trials (transition by group interaction, *P*=0.03). Finally, RL model fits showed only an increased value learning rate (*P*=0.01), but no changes in the influence of model-free or model-based action values on choice over learning (Figure 3h). Importantly, there were no significant session by group interactions between these patients and healthy volunteers for the stay probability analysis or RL model fits. Overall, these data suggest a different pattern of learning from experience in individuals with OCD, with a failure to learn the task-transition structure and exhibit model-based RL, but an increased influence of model-free action values.

### Explicit knowledge reduces model-free and increases model-based control

We next assessed how providing explicit information about the task structure changed behaviour, by comparing behaviour in sessions 3 and 4 in a group that received debriefing about task structure after session 3, and in another group that was not provided such information. To avoid ceiling effects in subjects who already acquired a model of the task, these analyses only included the 57 subjects for whom a likelihood ratio test indicated model-based RL was not being used significantly in session 3, as described above. Among these subjects, in session 4, more than 50% of those that were debriefed were identified by the likelihood-ratio test as using model-based RL (21/41), while in the absence of debriefing, only one subject became model-based (1/16; z=3.13, *P*=0.002, z-test for difference in proportions; Figure 4a, f). Consistently, debriefing strongly affected how events on each trial influenced the subsequent choice (Figure 4b, c, g, h), with increased influence of state transition (*P*<0.001; session by group interaction *P*=0.03; permutation tests) and transition-outcome interaction (*P*<0.001; session by group interaction *P*=0.01) on stay probability.

**Figure 4.**
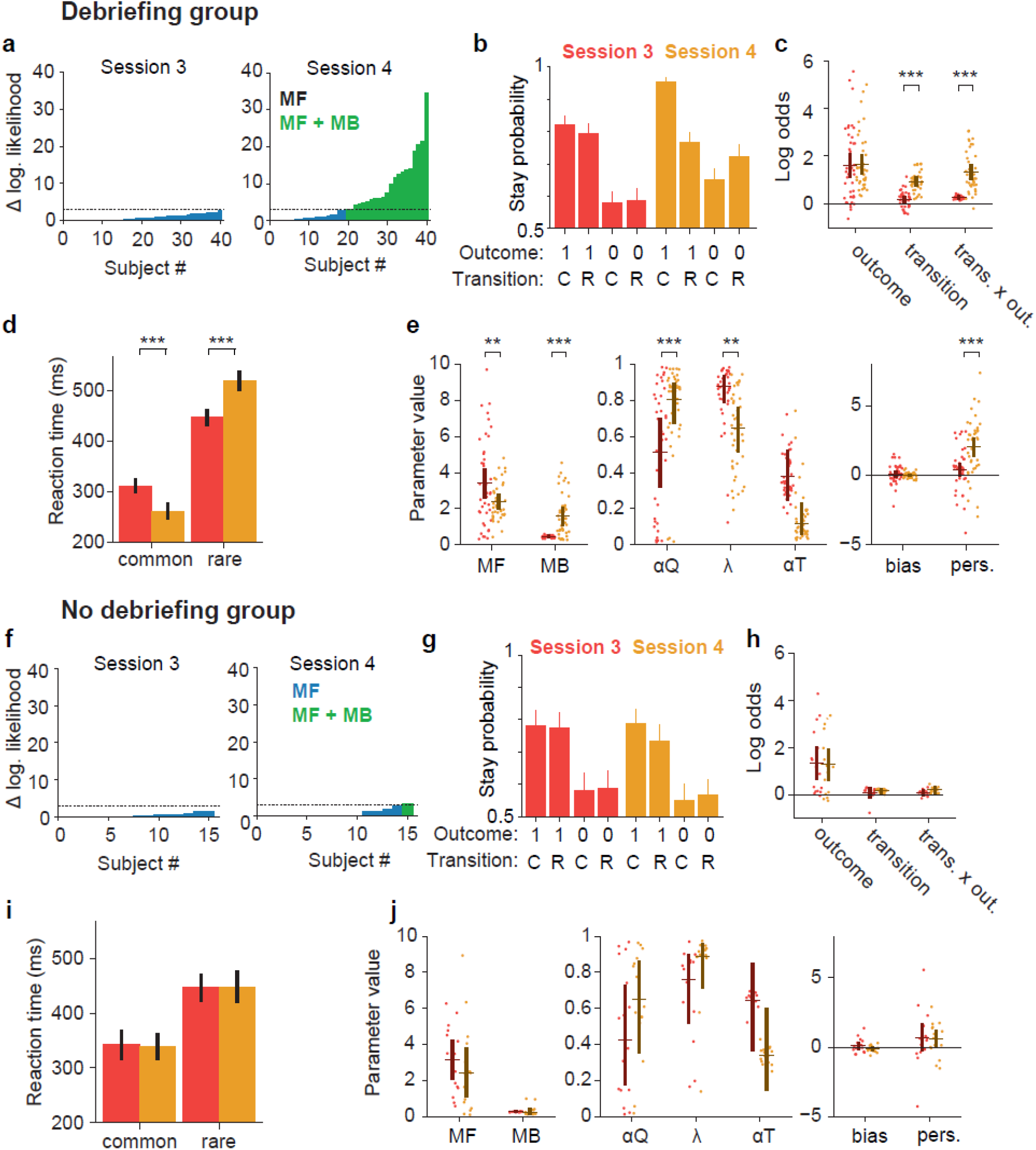
Explicit knowledge modifies the balance between model-based and model-free control. **(a, f)** Per-subject likelihood ratio test for use of model-based strategy on session 3 (left panel) and session 4 (right panel) among 57 healthy volunteers that did not use model-based RL significantly in session 3, as indexed by a likelihood ratio test. Data were analysed separately for groups with (**a**) and without (**f**) debriefing. Y-axis shows difference in log likelihood between a mixture (model-free and model-based) RL model and a model-free only RL model, with blue bars indicating subjects for whom the test favours the latter model, and green bars the subjects using a mixture model, using a p<0.05 threshold for rejecting the simpler model. **(b, g)** Stay probability analysis showing the probability of repeating the first step choice on the next trial as a function of trial outcome (rewarded or not rewarded) and state transition (common or rare). In these and remaining panels, red indicates session 3 (before instruction in the debriefing group) while yellow indicates session 4 (after instruction in the debriefing group). Error bars indicated the cross subject standard error of the mean (SEM). **(c, h)** Logistic regression analysis of how the outcome (rewarded or not), transition (common or rare) and their interaction, predict the probability of repeating the same choice on the subsequent trial. Dots indicate maximum a posteriori parameter values for individual subjects, while bars indicate the population mean and 95% confidence interval on the mean. **(d, i)** Cumulative distribution of reaction times for common and rare transitions. **(e, j)** Comparison of mixture model fits. Dots and bars are as in panel c. RL model parameters: MF, Model-free strength; MB, Model-based strength; αQ, Value learning rate; λ, Eligibility trace; αT, Transition probability learning rate; bias, Choice bias; pers., Choice perseveration.

RL mixture model fits of pre and post debriefing data (Figure 4e, j) confirmed that the influence of model-based action values on choice was increased by debriefing (*P*<0.001; session by group interaction *P*=0.01). Furthermore, the influence of model-free action values on choice reduced after debriefing (*P*=0.008), while value learning rates increased (*P*<0.001), though the session by group interactions were not significant (*P*=0.7 and *P*=0.6 respectively). In addition to modifying choice behaviour, debriefing increased differences in second-step key-press reaction times between common and rare transition trials (session-transition interaction *P*<0.0001, repeated measures ANOVA; Figure 4d), further supporting that the influence of state transition on RT in this task comprises both a motor component, which is independent of the use of model-based RL, and a cognitive component which manifests when subjects are using model-based RL. No significant differences were found for any comparisons between sessions 3 and 4 in the no debriefing group (Figure 4f-j).

Finally, among participants recruited in Lisbon, where neuropsychological data was available, we tested for correlations between test scores, namely from the Corsi block tapping test (assessing visuospatial working memory) and a Go/No-Go task (number of No-Go errors and reaction-time, assessing impulsivity), with several behavioural measures, specifically the outcome and transition-outcome interaction logistic regression predictor loadings, as well as the RL model parameters controlling the influence of model-free and model-based values on choice. Significant correlations were not found, neither among all healthy volunteers using data from session 3 (−0.27<r<0.31; 0.054<*P*<0.8), nor among the debriefing group using data from session 4 (−0.45<r<0.38; 0.07<*P*<0.8).

### Explicit knowledge affects model-free value updates and choice perseveration

Unexpectedly, the RL model also indicated that the eligibility trace parameter decreased after debriefing (*P*=0.004; session by group interaction *P*=0.03). This parameter controls the relative influence of the second-step state’s value and the trial outcome on updates to model-free first-step action values. Debriefing increased the influence of the second-step state value and decreased that of the trial outcome. As there is no obvious reason why providing task structure information should change model-free eligibility traces, we hypothesized that this effect is in fact mediated by the influence of task structural knowledge on representation of the task state space. By telling subjects that the reward probabilities depend on the second-step state reached, these states are likely made more distinct and salient in their internal representation of the task, and hence better able to accrue value, which can then drive model-free updates of first step action values. Consistent with this interpretation, subjects who, following debriefing, had large increases in the strength of model-based control, indicating that they had correctly understood the task structure, also had a larger decrease in the eligibility trace parameter (r=-0.34, *P*=0.03; Figure S3).

Debriefing also increased how often subjects repeated choices independent of subsequent trial events, as reflected by a significant increase in the ‘perseveration’ parameter of the RL model (*P*<0.001; session by group interaction *P*=0.001). This may result from information that reward probabilities on the left and right reversed only occasionally and are thus stable for extended periods of time. In this case, one would expect a reduction in perseveration across the course of each block, from shortly after a reversal, when reward probabilities are stable, to late in the block, when the next reversal is anticipated. Consistent with this hypothesis, we found that participants with larger post-debriefing increases in overall perseveration also had larger declines in perseveration within post-debriefing non-neutral blocks, from trials 10-20 (early) to 30-40 (late; r=-0.35, *P*=0.02; Figure S3).

Since all participants in the no debriefing group were recruited in Lisbon, while the debriefing group also included participants from the New York site, all analyses were repeated including only participants recruited in Lisbon, which did not affect the results qualitatively (data not shown). Furthermore, we tested for differences in learning or debriefing effects between the Lisbon and New York debriefing groups, and did not find any significant differences (Supplementary table sT1).

### Explicit knowledge increases model-based control in OCD

In the 41 of 46 individuals with OCD that were model-free at session 3, 37% (15 subjects) started using model-based RL after debriefing (Figure 5a, likelihood ratio test with threshold P=0.05). Consistent with this, the stay probability analysis showed increased loading on both the transition (*P*=0.002) and transition-outcome interaction (*P*<0.001) predictors, similarly to that observed in healthy controls (Figure 5b, c). Increased use of model-based RL after debriefing was confirmed by model fitting (Figure 5d), which showed increased influence of model-based action values on choice (*P*<0.001), and a trend towards reduced influence of model-free action values (*P*=0.06). As in healthy volunteers, debriefing in participants with OCD increased differences in second step reaction times between common and rare transition trials (session-transition interaction P<0.0001, repeated measures ANOVA; Figure 5d), while overall reaction times remained slower than in healthy volunteers (*P*=0.045, linear mixed model). Again similarly to healthy volunteers, debriefing reduced the value of the eligibility trace parameter (*P*=0.01), and this decrease correlated with increased use of model-based RL (r=-0.56, P=0.0001; figure S3). Though debriefing increased choice perseveration in OCD subjects (*P*=0.02), the effect was significantly smaller than in healthy volunteers (session by group interaction *P*=0.04), and unlike in healthy volunteers, did not correlate with decreases in perseveration from early to late in blocks after debriefing (r=-0.09, *P*=0.5; figure S3). This was the only significant interaction between debriefing and OCD diagnosis in direct comparisons with data from healthy volunteers for stay probability analysis and RL model fits.

**Figure 5.**
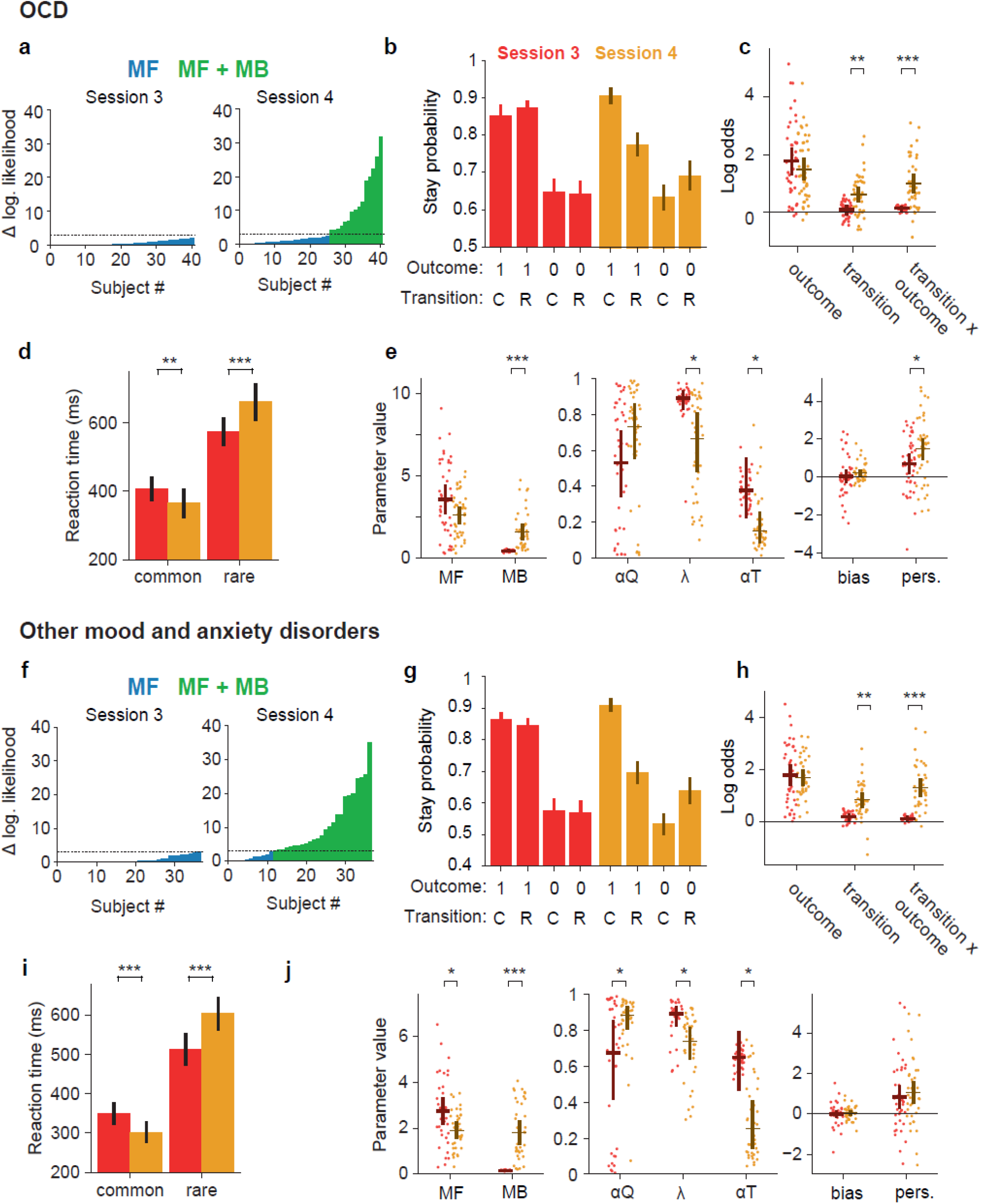
Explicit knowledge enhances model-based control in OCD. Among participants in the clinical groups, we analysed the effects of debriefing in 41 individuals with OCD (panels **a**-**e**) and 37 individuals with other mood and anxiety disorders (panels **f**-**j**) that did not use model-based RL in session 3. **(a, f)** Per-subject likelihood ratio test for use of model-based strategy on session 3 (left panel) and session 4 (right panel). Y-axis shows difference in log likelihood between mixture (model-free and model-based) RL model and model-free only RL model, with blue bars indicating subjects for whom the test favours the model-free only model, and green bars the subjects using a mixture model, using a p<0.05 threshold for rejecting the simpler model. **(b, g)** Stay probability analysis showing the probability of repeating the first step choice on the next trial as a function of trial outcome (rewarded or not rewarded) and state transition (common or rare). Error bars indicated the cross subject standard error of the mean (SEM). In this and all panels, red indicates session 3 (before instruction) while yellow indicates session 4 (after instruction). **(c, h)** Logistic regression analysis of how the outcome (rewarded or not), transition (common or rare) and their interaction, predict the probability of repeating the same choice on the subsequent trial. Dots indicate maximum *a posteriori* loading for individual subjects, bars indicate the population mean and 95% confidence interval on the mean. **(d, i)** Cumulative distribution of reaction times for common and rare transitions. **(e, j)** Comparison of mixture model fits. Dots and bars are as in panel c. RL model parameters: MF, Model-free strength; MB, Model-based strength; αQ, Value learning rate; λ, Eligibility trace; αT, Transition probability learning rate; bias, Choice bias; pers., Choice perseveration.

In the group with other mood and anxiety disorders, among 37 subjects that were model-free at session 3, debriefing increased the influence of transition (*P*=0.002) and transition-outcome interaction (*P*<0.001) predictors on stay probability (Figure 5f, g), similarly to healthy volunteers and individuals with OCD. The RL model fit confirmed that debriefing increased the influence of model-based action values on choice (*P*<0.001), and reduced influence of model-free action values (*P*=0.016; Figure 5h). As in the other groups, debriefing increased the difference in second-step reaction times between common and rare transition trials (*P*<0.0001), and there was no difference in reaction time effects between this group and healthy controls (*P*>0.3). Debriefing effects observed in healthy volunteers for the value learning rate and eligibility trace parameters were also replicated here (*P*=0.014 and *P*=0.04 respectively), with decrease in the latter again correlating with increased use of model-based RL (r=-0.45, P=0.005; figure S3). However, there was no effect of debriefing on choice perseveration (*P*=0.4; session by group interaction *P*=0.001), and no correlation across subjects between the debriefing effect on perseveration and post debriefing change in perseveration from early to late in blocks (r=-0.16, p=0.36). This was the only significant session by group interaction for stay probability analysis and RL model fits. Overall, this suggests that, with the exception of model-free strength, that was not significantly reduced by debriefing in individuals with OCD, both clinical groups were influenced by debriefing similarly to healthy volunteers, with all groups equally able to use the information about task structure to employ a model-based strategy.

To further explore potential effects of medication, we also tested for differences in RL strategies between the clinical groups recruited in Lisbon, the majority of whom were receiving pharmacological treatment (13/16 in OCD group, 14/16 in mood and anxiety disorders group), and the clinical groups recruited in New York, who were tested in the absence of such treatment. We found that debriefing reduced the strength of model-free RL and increased the value learning rate in treated, but not untreated, individuals with OCD (*P*=0.04 for group-debriefing interactions in both cases; Supplementary table sT2, Supplementary figure s4). Among individuals with other mood and anxiety disorders, significant differences were not found between Lisbon and New York samples (Supplementary table sT3).

Finally, among clinical groups recruited in Lisbon, where neuropsychological data was available, we tested correlations between test scores from the Corsi block tapping test or a Go/No-Go task, and outcome or transition-outcome interaction logistic regression predictor loadings, as well as model-free or model-based the RL model parameters, as described above for healthy volunteers. While in the OCD group we did not find significant correlations (data not shown), in the with mood and anxiety disorder group there were significant positive correlations between reaction time in the Go/No-Go task and several measures of model-based control, namely the transition-outcome interaction predictors from sessions 3 (r=0.69; P=0.007), and 4 (r=0.54; P=0.048), and the fitted model-based strength parameter value from session 4 (r=0.58; P=0.03). Other correlations were not significant.

## Discussion

We developed a simplified two-step task to examine how model-based and model-free RL contribute to behaviour in healthy and clinical populations, when task structure must be learned directly from experience. This allowed for subsequent testing of modifications of behavioural strategies once information about task structure was provided. Uninstructed behaviour was initially model-free, with strong direct reinforcement of choices by rewards from the start of the first session, but no evidence of subjects using knowledge of task structure early on. In fact, even after extensive experience, model-based control emerged only in a minority of subjects. This is striking given the relative simplicity of the task and suggests that humans are surprisingly poor at learning causal models from experience when they lack prior expectations about task structure. When learning from experience, individuals with OCD were impaired in their use of model-based control and biased towards a model-free strategy. Providing explicit information about task structure strongly increased the use of model-based control, across all tested populations, including individuals with OCD, with additional unexpected effects on model-free action value updates and choice perseveration. The absence of a model-based control deficit in OCD following debriefing is surprising, given the compelling evidence for such deficits in the original two-step task. This raises questions about task properties underlying the different pattern of deficits and suggests the possibility that real-world deficits in OCD patients may be rather due to an excessive use of a model-free learning system.

The increasing influence of model-based RL with experience contrasts with habit formation in rodent instrumental conditioning, where actions are initially goal-directed but become habitual with extended experience^44^, a process thought to involve a transition from model-based to model-free control^4^. This transition, and arbitration between model-based and model-free control more generally, has been proposed to occur through meta-cognitive mechanisms which assess whether the benefits of improved prediction accuracy are worth the costs of model-based evaluation^4,12,13^. The different trajectory in the current task likely results from a more complex state space that increases model uncertainty in early learning and makes model-based learning more demanding, and from ongoing changes in reward probability that prevent the model-free system from converging to accurate value estimates in late learning^4^. In fact, it has been recently suggested that performance during initial stages of action selection tasks may be primarily based on trial-and-error exploration, with progression towards model-based RL occurring in intermediate stages, as subjects acquire a model of the environment^45^.

Our finding that model-free RL dominates uninstructed behaviour on a two-step task contrasts with recent arguments from Silva & Hare^32^ that humans are primarily model-based learners on two-step tasks, and that apparent model-free behaviour is in fact model-based control using muddled or incorrect task models. We cannot rule out the possibility that some apparently model-free behaviour at later uninstructed sessions in our task was in fact model-based control with an incorrect model. However, we do not think this is a plausible overall explanation for the observed predominance of model-free behaviour prior to instructions. Firstly, because stay probabilities at session one showed a strong main effect of outcome but no transition-outcome interaction, i.e. the canonical picture of a model-free agent. This is not consistent with Silva and Hare’s simulations of agents with muddled models, which show a strong effect of transition-outcome interaction^32^. Secondly, our subjects showed a direct reinforcing effect of reward on first-step choice from their very first interactions with the task. It does not appear likely that subjects almost instantly acquire muddled models of the task which happen to produce the exact effect predicted by model-free reinforcement. Rather, we propose that, consistent with findings as early as Thorndike’s law of effect^46^, rewards in our task had a direct reinforcing effect on actions performed shortly prior to their being obtained. While these findings provide evidence that subjects use model-free control in unfamiliar domains, this speaks only indirectly to the question of whether model-free RL or muddled models underlie apparent model-free behaviour in the original two-step task.

Providing explicit information about task structure strongly boosted the influence of model-based RL and reduced the influence of model-free RL. This complements Silva and Hare’s findings that model-based control is increased by making instructions more complete and embedding them in a narrative to make them easier to remember and understand. Such instruction effects are consistent with meta-cognitive cost-benefit decision making, since an accurate model of the task structure will boost the estimated accuracy of model-based predictions and hence the expected payoffs from model-based control. Our findings also build on extensive literature examining how instruction and experience interact to determine human behaviour, in tasks that do not discriminate model-free and model-based control. Early work examining instruction effects on operant conditioning found that after explicit information about the schedule of reinforcement, responses match the contingencies explained to subjects (e.g. fixed interval, variable interval or fixed ratio), even when these differ substantially from the actual contingencies^33–35^. In common with our study, these results emphasize that humans learn about task structure much more readily from explicit information than via trial and error learning. More recent work has focused on the effect of advice, i.e. informing subjects that one option is particularly good or bad, on reward guided decision making in probabilistic settings^38,39^. Such advice impacts not only initial estimates of how good or bad different options are, but also modifies subsequent learning, by up-weighting and down-weighting outcomes. Whether such bias effects extend to learning about task structure, in addition to simple reward learning, is an open question for further work. Functional neuroimaging has also shown that instructions change responses to outcomes in the striatum, ventromedial prefrontal cortex and orbitofrontal cortex, potentially mediated by representations of instructed knowledge in the dorsolateral prefrontal cortex^37,40,47^. Our task provides a potential tool for extending such mechanistic investigation of instruction effects into the domain of task structure learning and model-based RL.

Our findings may have translational relevance for OCD. Prior studies have shown that individuals with OCD, as well as healthy volunteers with self-reported OCD-like symptoms, have deficits in model-based control in the original two-step task^28,30^. There is also data showing that these findings reflect a transdiagnostic compulsivity dimension, rather than an OCD-specific characteristic^30,48^. Consistent with these reports, comparing OCD subjects with healthy volunteers we found evidence for impaired acquisition of model-based control, and increased influence of model-free values, when learning directly and exclusively from experience. No difference was found in comparisons between healthy volunteers and individuals with other mood and anxiety disorders during uninstructed experience. Surprisingly, following debriefing we did not observe deficits in the ability of OCD subjects to adopt a model-based strategy, demonstrating that, under some conditions, individuals with OCD recruit model-based control as readily as healthy volunteers, which is of particular interest given the established efficacy of cognitive-behavioural therapy (CBT) in the treatment of OCD^49^. The evidence for increased influence of model-free values prior to instructions rather suggests that in OCD there may be a bias towards increased use of model-free RL, which is consistent with the view that OCD may be driven by an hyperactive habit system^28,50,51^. Potentially relevant regarding this hypothesis, while debriefing had no effect on use of model-free RL in non-medicated patients, this was reduced after debriefing in treated patients, with learning rates increased in medicated but not unmedicated participants with OCD. Treated OCD subjects were thus, in these respects, closer to healthy subjects. Although it has been shown that, in OCD subjects, cognitive-behavioural therapy does not change use of model-based control in the original two-step task^52^, our results suggest that pharmacological treatment may have an effect on the ability to suppress model-free control and modify behaviour once a correct model is acquired. We did make a trend observation that the influence of model-free values on choices increases with experience in OCD, but not in healthy volunteers (group difference: P = 0.07). Given the clinical relevance of this observation, it would be valuable to attempt a formal replication, potentially using more extensive training in studies including clinical samples.

There are substantial differences between our paradigm and the original two-step task, that may explain the different pattern of deficits observed in individuals with OCD. Our task is structurally simpler due to having no choice at the second-step, which will reduce working memory load and hence make model-based control easier. Indeed, while in the original two-step task working memory capacity is correlated with the use of a model-based strategy^19,20^, we did not find any such correlations here, neither in healthy volunteers nor in clinical populations. The fact that subjects in our task have extensive prior experience before being told its structure may also help them understand or remember this information when it is provided. Furthermore, in our task, actions and states were differentiated by location rather than identity of visual stimuli, and these locations were fixed across trials rather than randomized as in the original two-step task. This allows model-free RL operating over spatial-motor representations, as has been demonstrated by others^53^, to contribute more meaningfully to choice. Fixed spatial-motor contingencies also permit of use of action-outcome, rather than stimulus-outcome, mappings for model-based control, with the former thought to preferentially recruit the anterior cingulate cortex, rather than the OFC^54,55^. They may also underpin our observation of reaction-time differences following common and rare transitions, even when choices were model-free, that is consistent with a recent report that motor actions are distinct according to state transitions, even when choice is model-free^56^. Finally, unlike the original two-step task, where model-based and model-free RL achieved similar reward rates^42,43^, here use of model-based RL positively correlated with reward rate, generating a desirable trade-off between performance and cognitive effort that may influence arbitration between strategies^43^.

In addition to increasing model-based control, debriefing had unexpected effects on model-free value updates, increasing the influence of second-step state values relative to trial outcomes on model-free first-step action values. This effect was robust and replicated in both clinical groups and healthy volunteers. We hypothesized that this was mediated by effects of debriefing to modify internal representations of the task state-space. Knowledge that the reward probability depended on the second-step state that was reached likely made internal representation of these states more salient and differentiated, and hence better able to accrue value, and thus driving model-free learning at the first step. Consistent with our hypothesis, subjects who became more strongly model-based following debriefing - indicating that they had acquired a correct model of the task, showed larger changes in model-free value updates. This result emphasizes that model-free RL operates over an internal representation of the states of the external world that must be learnt from sometimes ambiguous experience and is malleable in the face of new information^57^. A second unexpected effect of debriefing was an increased tendency to repeat choices, as indexed by the RL model’s perseveration parameter. We hypothesized that this effect was mediated by explicit knowledge that the reward probabilities changed only occasionally. Consistent with this hypothesis, post-debriefing increases in perseveration correlated with decreases in perseveration over the course of each block. It thus seems that, after debriefing, subjects inferred the occurrence of a reward probability reversal, and expected stability in the trials immediately following. These two unexpected findings show that the ‘model’ of a task that may be acquired through explicit information comprises not just the action-state contingencies that are required for model-based RL, but also beliefs about which distinct states of the environment are relevant for behaviour, and how the world may change over time, both of which can influence ‘model-free’ value learning. It is also important to note that the most significant difference between clinical populations and healthy volunteers following debriefing was that, while the latter became more perseverative in their choices, this effect was smaller in OCD, absent in individuals with other mood and anxiety disorders, and did not correlate in either patient population with changes in perseveration from early to late in post-debriefing blocks. This evidence that inference based updating was impaired in OCD and other psychiatric diagnoses is particularly interesting given that the orbitofrontal cortex, which is consistently dysfunctional in OCD patients^58–60^, is thought to build cognitive maps needed to infer task states that are not directly observable from sensory input^61^.

We should note a number of limitations and directions for future studies. First, as well as standard blocks, in which transition probabilities were asymmetric, the task included neutral blocks with equal reward probabilities on the left and right sides. For comparison with the original two-step task, it would be interesting to assess the influence of the latter on the dominance of model-free control in the uninstructed case. Equally, it would be worth looking parametrically at the effect of instructions as a function of the amount of uninstructed experience in our task. Second, since prior work has identified that symptom dimensions can be a better predictor of behavioural phenotypes than clinical diagnoses, applying our task with dimensional methods in large online samples could provide further insight into clinical differences in learning and instruction effects^48^. Finally, although we know of no study showing a positive correlation between years of education and use of model-based or model-free control, the small but significant difference in terms of education between healthy volunteers and individuals with the mood and anxiety disorders might limit the comparisons between these samples. We did observe substantial heterogeneity across subjects in uninstructed behaviour, and it is likely that increased variability is an inherent feature of uninstructed tasks that may complicate assessing group differences.

In summary, we developed a new sequential decision task which dissociates the effects of uninstructed experience and explicit information on RL strategy. We found that model-free RL dominates initial behaviour and maintains a strong influence throughout uninstructed learning, with model-based RL emerging only in a subset of individuals prior to receiving task structure information, and to a lesser extent in individuals with OCD. Receiving such information strongly increased model-based control, both in healthy individuals and those with OCD and other mood and anxiety diagnoses. Use of this new task to dissociate effects of implicit and explicit information on RL strategy thus offers further insight into the content of learning and the imbalance between RL systems in neuropsychiatric disorders.

## Methods

### Participants and Testing Procedures

The research protocol was conducted in accordance with the declaration of Helsinki for human studies of the World Medical Association and approved by the Ethics Committees of the Champalimaud Centre for the Unknown, NOVA Medical School and Centro Hospitalar Psiquiátrico de Lisboa (CHPL), and the Institutional Review Board of the New York State Psychiatric Institute (NYSPI). Written informed consent was obtained from all participants. Clinical samples were recruited at the Champalimaud Clinical Centre (CCC), CHPL and the NYSPI. In each of these centres, individuals with OCD were recruited from clinical or research databases. A mood and anxiety disorder control group was recruited randomly from patient lists (CCC and CHPL), or sequentially (NYSPI), among individuals with the following diagnoses: major depressive episode or disorder, dysthymia, bipolar disorder, generalized anxiety disorder, post-traumatic stress disorder, panic disorder or social anxiety disorder. Healthy controls were recruited sequentially as a convenience sample of community-dwelling participants and tested at the same locations.

Following consent, each participant was screened for the presence of exclusion criteria using a clinical questionnaire assessing history of: acute medical illness; active neurological illness; clinically significant focal structural lesion of the central nervous system; history of chronic psychosis, dementia, developmental disorders with low intelligence quotient or any other form of cognitive impairment and illiteracy. Active psychiatric illness, including substance abuse or dependence, was also an exclusion criterion, with the exception of the diagnoses defining inclusion in the OCD and the mood and anxiety groups. In the absence of exclusion criteria, each participant then performed the simplified two-step task (see below).

Participants also performed a battery of structured interviews, scales and self-report inventories, including the MINI Neuropsychiatric Interview^62^, the Structured Clinical Interview for the DSM-IV^63^, the Yale-Brown Obsessive-Compulsive Scale (Y-BOCS)^64,65^ and the State-Trait Anxiety Inventory (STAI)^66^. As the groups recruited in Lisbon were assessed using the Y-BOCS-II while the groups recruited in New York were assessed using the original Y-BOCS, we converted the Y-BOCS-II score into original Y-BOCS score by transforming each item which was scored as 6 into a score of 5^65,67^. In the groups recruited in Lisbon, the Beck Depression Inventory-II (BDI-II)^68^ was also applied to assess depressive symptoms, the Corsi block-tapping task to assess working memory^69^ and a Go/No-Go task to assess impulsivity^70^, while in New York, the Depression Anxiety Stress Scales (DASS)^71^ were applied to assess symptoms of depression, anxiety and stress. Group differences in sociodemographic and psychometric measures were tested using one-way ANOVA for continuous variables (with Tukey’s HSD for multiple comparisons) and Pearson’s chi-squared for categorical variables. Correlations between neuropsychological test measures (Corsi; Go/No-GO) and the simplified two-step task measures were performed using Pearson’s product moment correlation coefficient.

### Simplified two-step task

The simplified two-step task was implemented in MATLAB R2014b using Psychtoolbox (Mathworks, Inc., Natick, Massachusetts, USA). The task consisted of a self-paced computer interface with 4 circles always visible on the screen: 2 central circles (upper and lower) flanked by two side circles (left and right) (Figure 1). Each circle was coloured yellow when available for selection, and black when unavailable, and could be selected by pressing the corresponding arrow key (up, down, left or right) on the computer keyboard. Each trial started with both of the central circles turning yellow, prompting a choice between the two (Figure 1a). This first step choice then activated one of the side circles in a probabilistic fashion, according to a structure of transition probabilities described below (Figure 1b). The active side circle could be selected with the corresponding arrow key, resulting either in reward (indicated by the circle changing to the image of a coin) or no reward (indicated by the circle changing to black). The reward probabilities on the right and left side changed in blocks that were either neutral (p=0.4 on each side) or non-neutral (p=0.8 on one side and p=0.2 on the other; Figure 1c). Changes from non-neutral blocks were triggered based on each subject’s behaviour, occurring 20 trials after an exponential moving average (tau = 8 trials) crossed a 75% correct threshold. In half of the cases this led to the other non-neutral block (reward probability reversals), and the other half to a neutral block. Changes from neutral blocks occurred with 10% probability on each trial after the 40^th^ trial of that block, and always led to the non-neutral block that did not precede that neutral block. All participants performed 1200 trials on the same day, divided in 4 sessions of 300 trials each.

We ran two variants of the task which differed with respect to whether the transition probabilities linking the first-step actions to the second step states were fixed or underwent reversals. In both cases these probabilities were defined such that choosing one of the central circles (e.g. high) would cause one of the side circles (e.g. left) to turn yellow with high probability (p=0.8 – common transition), while causing the other side circle to turn yellow only in a minority of trials, i.e., with low probability (p=0.2 – rare transition). Choosing the other central circle would lead to common and rare transitions to the opposite sides. In the Fixed task, the transition probabilities were fixed for each individual throughout the entire task (e.g., common transitions for high-left and low-right, and rare transitions for high-right and low-left). In the Changing task, the transition probabilities underwent reversals on 50% of reward probability block changes after non-neutral blocks, such that the common transition became rare and vice versa (Figure 1b). In an initial group of healthy volunteers recruited in Lisbon, subjects were randomized between the two versions of the task. In all clinical samples as well as healthy volunteers from New York, however, only the Fixed task was used.

Prior to starting the task, subjects were given minimal information about task structure. They were only told that arrow keys could be used to interact with the screen, and that the image of a coin signalled accrual of a monetary reward. To test how providing explicit information about the task structure affected behaviour, debriefing was provided between the 3rd and the 4th sessions in some participants, with the 4th session of the task performed immediately after debriefing. Among healthy volunteers recruited in Lisbon and randomized between the two versions of the task, debriefing was performed in 17 participants performing the Fixed version and in 16 participants performing the Changing version of the task. In all other samples, debriefing was performed for everyone. Please see supplementary material for the specific information provided to subjects prior to the task and during debriefing.

### Data analysis

Data analysis was performed using Python (Python Software Foundation, http://python.org) and SPSS (Version 21.0, SPSS Inc., Chicago, IL, USA), and centred on three main analyses of behaviour on the task: reversal analysis, logistic regression analyses of stay probability, and RL model comparison and fitting. The reversal analysis assessed overall task performance according to the average first step choice trajectory around reward probability reversals between non-neutral blocks, from which we extracted two measures. One was the fraction of correct choices at the end of the block before the reversal, with correct defined as the first step choice with a common transition to the side (i.e., state), with highest reward probability. The second measure was the time constant of adaptation to the reversal, estimated by a least squares exponential fit to the cross subject mean choice trajectory following the reversal. For the Changing version of the task, reversal analysis was performed according to the average trajectory for reversals in both transition and reward probabilities. Importantly, while reversal analyses provide information about how well subjects are able to track which option is correct, they do not differentiate between use of model-free and model-based strategies.

The first analysis used to assess model-free vs. model-based behavioural strategies was an analysis of ‘stay-probability’^11,16,25,26,17–24^, defined as the probability of repeating the first-step choice on any given trial as a function of the outcome (rewarded or not) and transition (common or rare) on the previous trial. In addition to plotting raw stay probabilities, we analysed the effect of trial events on the subsequent choice using a logistic regression model with several binary predictors. The *outcome, transition* and *transition-outcome interaction* predictors modelled the influence of the previous trial’s outcome, transition and their interaction on the probability of repeating the previous first step choice. We additionally included a *bias* predictor capturing bias towards the upper or lower circle, and a *correct* predictor, which modelled the influence of whether the previous trials choice was correct (i.e. high reward probability) on the probability of repeating that choice. The latter prevents spurious loading on the *transition-outcome interaction* predictor, which has been described in two-step tasks with high contrast between good and bad options, due to correlation between action values at the start of the trial and subsequent trial events^42^.

### RL modelling

Additional analyses of behavioural strategy were obtained by fitting reinforcement learning models to observed behaviour. We first detail the model used for the main analyses then a set of alternative models that were rejected by model-comparison. The model followed those typically used in analysis of the original two-step task^11^ in combining a model-based and a model-free RL component, both with value estimates contributing to behaviour. The model-free component maintained estimates of the values *Q*^*mf*^(*a*) of the first-step actions (up or down), and *V*(*s*) of the second step states (left and right). These values were updated as:

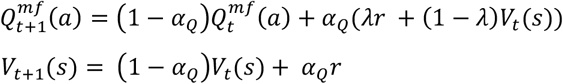

Where *r* is the reward obtained on trial *t* (1 or 0), *α*_*Q*_is the value learning rate and *λ* is the eligibility trace parameter.

The model-based component maintained estimates of the transition probabilities linking the first step actions to the second step states (*P*(*s*_2_|*a*_1_)), updated as:

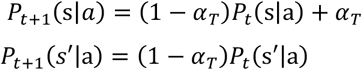

where *α*_*T*_ is a learning rate for transition probabilities, s is the second step state reached and *s*′ the second step state not reached on trial *t*.

At the start of each trial, model-based action values were calculated as:

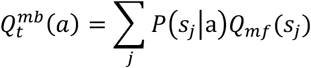

Model-free and model-based action values were combined with perseveration and bias to given net action values, calculated as:

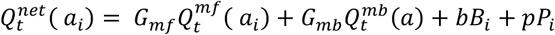

Where *G*_*mf*_ and *G*_*mb*_ are parameters controlling, respectively, the strength of influence of model-free and model-based action values on choice, *b* is a parameter controlling the strength of choice bias, *B*_*i*_ is a variable which takes a value of 1 for the high action and zero for the low action, *p* is a parameter controlling the strength of choice perseveration, *P*_*i*_ is a variable which takes a value of 1 if action *a*_*i*_ was chosen on the previous trial and 0 if it was not.

The model’s probability of choosing action *a*_*i*_ was given by 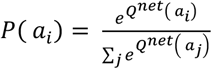.

For model comparison, several reduced variants were considered. For the *Model-free only* variant the model-based component was removed such that the net action values were:

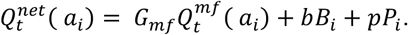

For the *Model-based only* variant the model-free component was removed such that the net action values were:

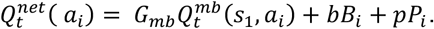

For the *No bias* variant the bias strength variable *b* was set to zero. For the *No perseveration* variant the perseveration strength variable *p* was set to zero.

### Hierarchical modelling

Fits of both the logistic regression model and reinforcement learning models to populations of subjects used a Bayesian hierarchical modelling framework ^72^, in which parameter vectors ***h***_*i*_ for individual sessions were assumed to be drawn from Gaussian distributions at the population level with means and variance ***θ*** = {***μ, Σ***}. The population level prior distributions were fit to their maximum likelihood estimate:

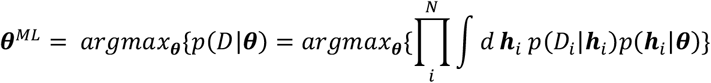

Optimization was performed using the Expectation-Maximization algorithm with a Laplace approximation for the E-step at the k-th iteration given by:

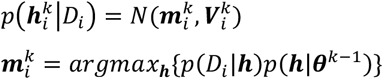

Where 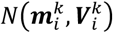 is a normal distribution with mean 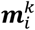 given by the maximum a posteriori value of the session parameter vector ***h***_*i*_ given the population level means and variance ***θ***^***k***−1^, and the covariance 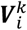 given by the inverse Hessian of the likelihood around 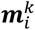. For simplicity we assumed that the population level covariance ***Σ*** had zero off-diagonal terms. For the k-th M-step of the EM algorithm the population level prior distribution parameters ***θ*** = {***μ, Σ***} are updated as:

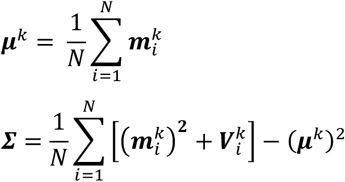

Parameters were transformed before inference to enforce constraints:

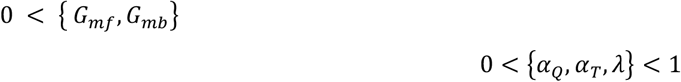

95% confidence intervals on population means ***μ*** were calculated as 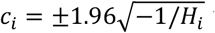 where *c*_*i*_ is the confidence interval for parameter *i* and *H*_*i*_ is the *i*-th diagonal element of the Hessian at ***θ***^*ML*^ with respect to ***μ***.

### Model comparison

To compare the goodness of fit for hierarchical models with different numbers of parameters we used the integrated Bayes Information Criterion (iBIC) score. The iBIC score is related to the model log likelihood *p*(*D*|*M*) as:

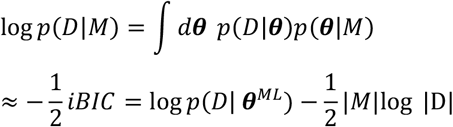

Where |M| is the number of fitted parameters of the prior, |D| is the number of data points (total choices made by all subjects) and iBIC is the integrated BIC score. The log data likelihood given maximum likelihood parameters for the prior log *p*(*D*| ***θ***^*ML*^) is calculated by integrating out the individual session parameters:

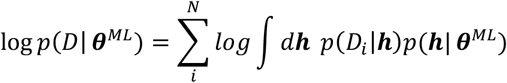

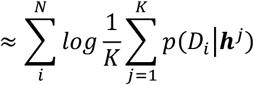

Where the integral is approximated as the average over K samples drawn from the prior *p*(***h***|***θ***^*ML*^).

### Permutation testing

Permutation testing was used to assess the statistical significance of learning and instruction effects on RL and logistic regression model fits, and the reversal analysis. To assess the effects of experience in the task we compared behavior between sessions 1 and 3, while to assess the effects of explicit knowledge we compared behavior between sessions 3 and 4 in the groups that did and did not receive instruction between these sessions.

To test for a significant difference in behavioral parameter *x* between conditions (e.g. session 1 vs 3), we evaluated the population mean value of the parameter for each conditions and calculated the difference Δx_*true*_ between them. We then constructed an ensemble of 5000 permuted datasets in which the assigments of sessions to the two conditions was randomised. Randomisation was performed within subject, such that the number of sessions from each subject in each condition was preserved. For each permuted dataset we re-ran the analysis and evaluated the difference in parameter *x* between the two conditions, to give a distribution of Δx_*perm*_, which in the limit of many permutations is the distribution of Δx under the null hypothesis that there is no difference between the conditions. The two tailed P value for the observed difference is given by:

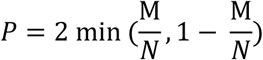

Where N is the number of permutations and M is the number of permutations for which Δx_*perm*_ > Δx_*true*_.

To assess significant differences in learning or debriefing effects between clinical groups and healthy controls, and for differences in the healthy controls between groups who did and did not receive debriefing, we tested for a significant interaction between session number and group. The significance of the interaction was assessed using a permutation test in which we evaluated the difference 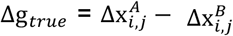 where 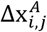 is the difference in behavioural parameter *x* between sessions *i* and *j* in group *A*, and 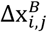 is the difference in behavioural parameter *x* between sessions *i* and *j* in group B. We then constructed an ensemble of 5000 permuted datasets by randomly permuting subjects between groups while preserving the total number of subjects in each group. We assessed 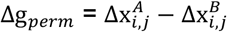 for each permuted dataset and calculated P values for the interaction as above.

## Data Availability

N/A

## Supplementary results

### Changing transition probabilities inhibit model-based control

In 42 healthy volunteers recruited in Lisbon, we tested a version of the task in which transition probabilities linking the first step actions and second-step states underwent periodic reversals. In this Changing version of the task (Figure S5), subjects were still able to successfully track which first step action was correct, choosing the correct option at the end of block well above chance level, (session 1: 0.77, session 3: 0.76; Figure S5c), and adapting to reversals in reward probabilities with a similar trajectory relative to the Fixed task (exponential fit tau, session 1: 22.8 trials, session 3: 14.0 trials). Again, neither of these measures of overall performance changed significantly between sessions 1 and 3 (P>0.3).

However, unlike in the Fixed task, the stay-probability analysis did not show increased influence of transition and transition-outcome interaction in session 3 relative to session 1 (*P*=0.2 for both), consistent with no changes in the use of model-based RL. On the contrary, there was an increase in the influence of trial outcome (*P*=0.01), associated with a model-free direct reinforcement strategy^11,42^ (Figure 51a,b). Similarly to the Fixed task, there was a significant correlation between loading on the transition–outcome interaction parameter and the number of rewards obtained (rho=0.4, *P*<0.01). Model comparison indicated that the mixture and model-free-only RL models fitted the data much better than the model-based-only model, and that the difference in BIC scores between the mixture model and model-free-only model was negligible (ΔiBIC = 3; Figure S5d left panel). Again similarly to the Fixed task, the model including both “bias” and “perseveration” parameters fits the data better than a model lacking these parameters (Figure S5d, right panel). For consistency with analysis of the Fixed task, we used the mixture model to look for differences in behaviour between sessions 1 and 3 but found no significant change in model parameters (Figure S5e). These data suggest that changes in the action-state transition probabilities prevented most subjects from learning a model-based strategy.

In this version of the task, debriefing did not increase the use of model-based RL among subjects for whom a likelihood ratio test indicated model-based RL was not being used significantly in session 3 (n=36; Figure S6). The fraction of subjects identified as using a model-based strategy at session 4 was the same in the debriefing and no-debriefing groups (debriefing group 2/12, no-debriefing group 4/24; z = 0, *P*=1, z-test for difference of proportions; Figure S2A, F). Subjects in the debriefing group adapted faster to reversals in session 4 than session 3 (*P*=0.03, Figure S6b), and the logistic regression analysis showed an increased influence of the trial outcome on subsequent choice in session 4 compared to 3 in the debriefing group (*P*=0.02, Figure S6d), but the session by group interaction did not reach significance in both cases (*P*=0.2 and P=0.06 respectively). The influence of the transition and transition-outcome interaction parameters on subsequent choice were unaffected by debriefing (P=0.99 and P=0.3 respectively, Figure S6d) and no parameters of the RL model differed significantly pre and post-debriefing (P>0.19, Figure S6e). As expected, no significant differences were observed in any analyses between sessions 3 and 4 in the no-debriefing group (Figure S6g-j). These results indicate that in the more complex Changing task, subjects either did not understand the debriefing or decided the effort of trying to use information about the task structure was not worthwhile.

**Supplementary table sT1.** Differences in learning and debriefing effects in healthy volunteers between the Lisbon and New York samples.

| Model parameters | Learning effects<sup>a</sup> | Debriefing effects<sup>a</sup> |
| --- | --- | --- |
| Model-free strength (MF) | 0.96 | 0.92 |
| Model-based strength (MB) | 0.26 | 0.25 |
| Value learning rate ($\alpha_Q$) | 0.37 | 0.56 |
| Eligibility trace ($\lambda$) | 0.2 | 0.1 |
| Transition learning rate ($\alpha_T$) | 0.14 | 0.62 |
| Choice bias | 0.4 | 0.73 |
| Choice perseveration | 0.69 | 0.32 |

<sup>a</sup>Permutation tests (5000 permutations) were used to assess differences in the fitted model parameter loadings between Fixed task Lisbon (n=40) and New York (n=27) samples in the effect of learning (defined as change between session 1 and 3) and debriefing (defined as change between session 3 and 4, taking only subjects who are MF at session 3). P-values for interactions between group and the effect of interest are shown.

**Supplementary table sT2.** Differences in learning and debriefing effects in individuals with OCD between the Lisbon and New York samples.

| Model parameters | Learning effects<sup>a</sup> | Debriefing effects<sup>a</sup> |
| --- | --- | --- |
| Model-free strength (MF) | 0.38 | **0.04** |
| Model-based strength (MB) | 0.1 | 0.66 |
| Value learning rate ($\alpha_Q$) | 0.88 | **0.04** |
| Eligibility trace ($\lambda$) | 0.87 | 0.87 |
| Transition learning rate ($\alpha_T$) | 0.95 | 0.24 |
| Choice bias | 0.86 | 0.58 |
| Choice perseveration | 0.17 | 0.79 |

<sup>a</sup>Permutation tests (5000 permutations) were used to assess differences in the fitted model parameter loadings between Fixed task Lisbon (n=16) and New York (n=30) samples in the effect of learning (defined as change between session 1 and 3) and debriefing (defined as change between session 3 and 4, taking only subjects who are MF at session 3). P-values for interactions between group and the effect of interest are shown.

**Supplementary table sT3.**
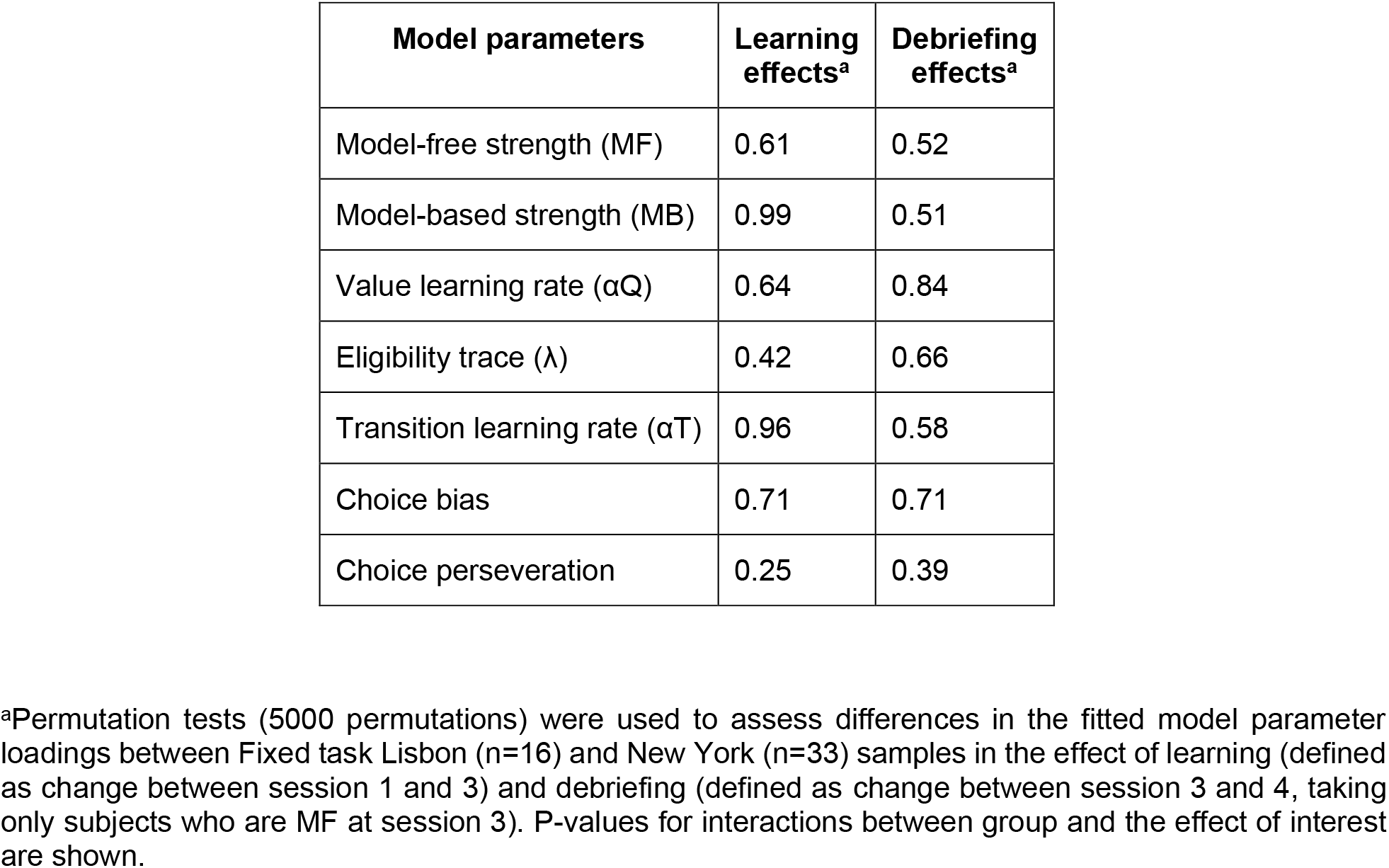
Differences in learning and debriefing effects in individuals with other mood and anxiety disorders between the Lisbon and New York samples.

**Supplementary figure S1.**
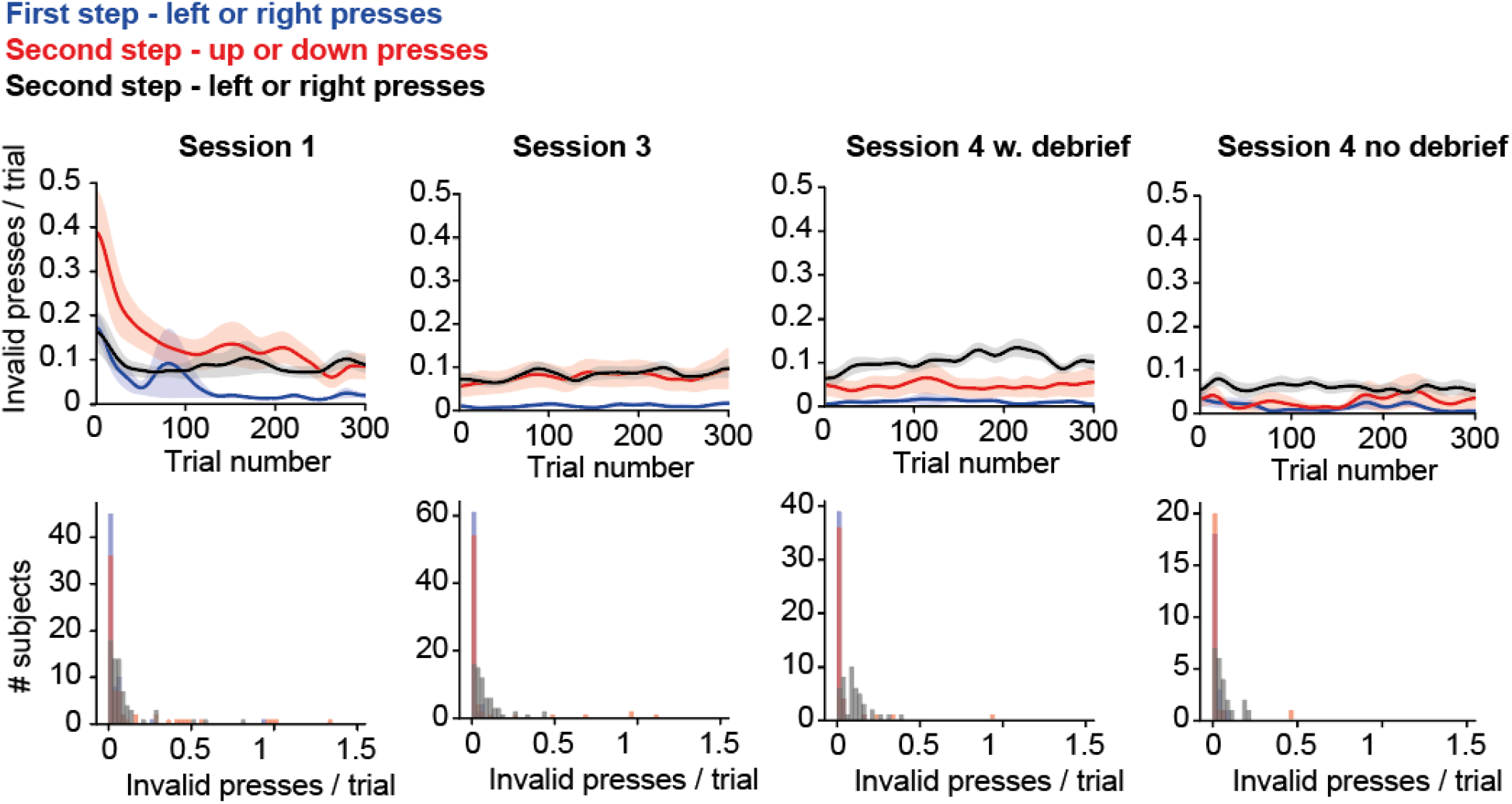
Invalid key presses. Top panels show the mean number of invalid key presses per trial as a function of trial number, Gaussian smoothed with an SD of 10 trials, shaded area shows SEM across subjects. Bottom panels show histogram of the mean number of invalid presses per trial across the entire session for each subject.

**Supplementary figure S2.**
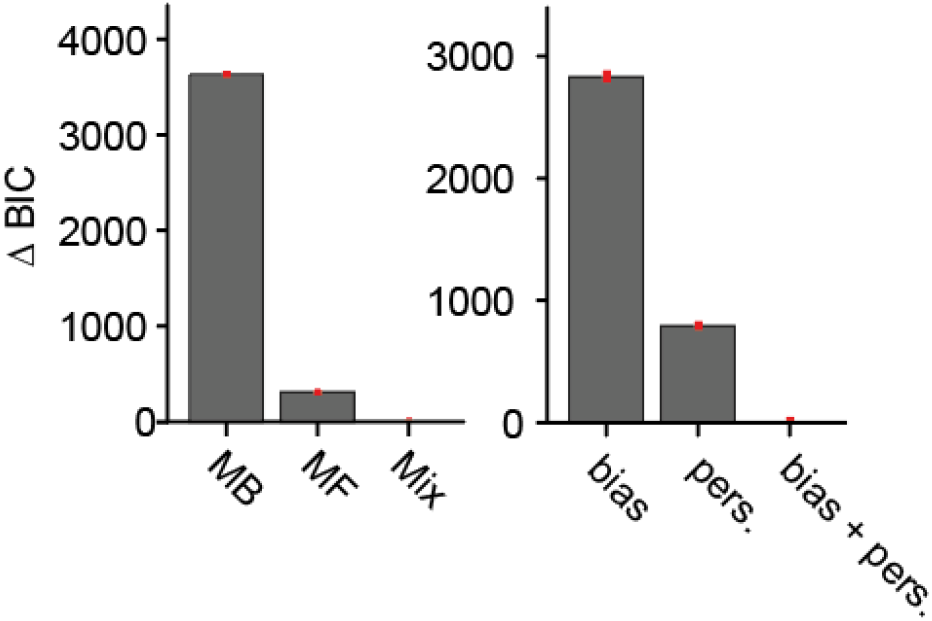
Model comparison. Model comparison for sessions 1-3 of the Fixed task. Panels show the difference in BIC score relative to the best fitting model. Left panel, comparison of model-based (MB), model-free (MF) and MB-MF mixture (Mix) models. Right panel, comparison of mixture model with bias parameter, perseveration parameter, and bias + perseveration parameters.

**Supplementary figure S3.**
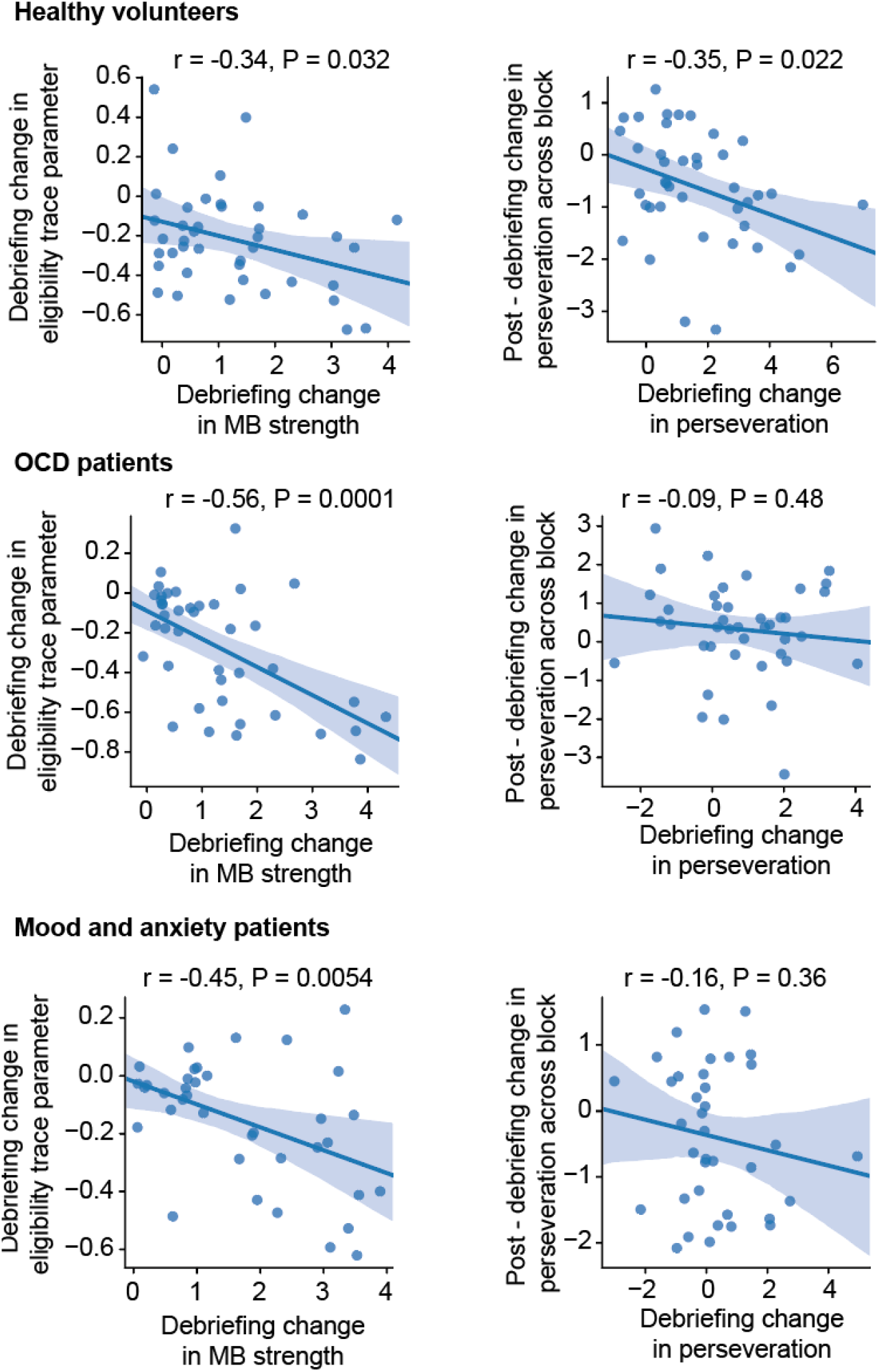
Debriefing effect correlations. Left panels – Correlation across subjects between the effect of debriefing on subjects use of model-based RL (as assessed by the RL model’s model-based weight parameter) and that on the RL model’s eligibility trace parameter. Right panels – Correlation between the effect of debriefing on subjects overall perseveration (as assessed by the RL models perseveration parameter), and post debriefing, their change in perseveration from early to late in blocks, assessed using logistic regression analysis of data from early (10-20 trials post block transition) and late (30-40 trials) in each block.

**Supplementary figure S4.**
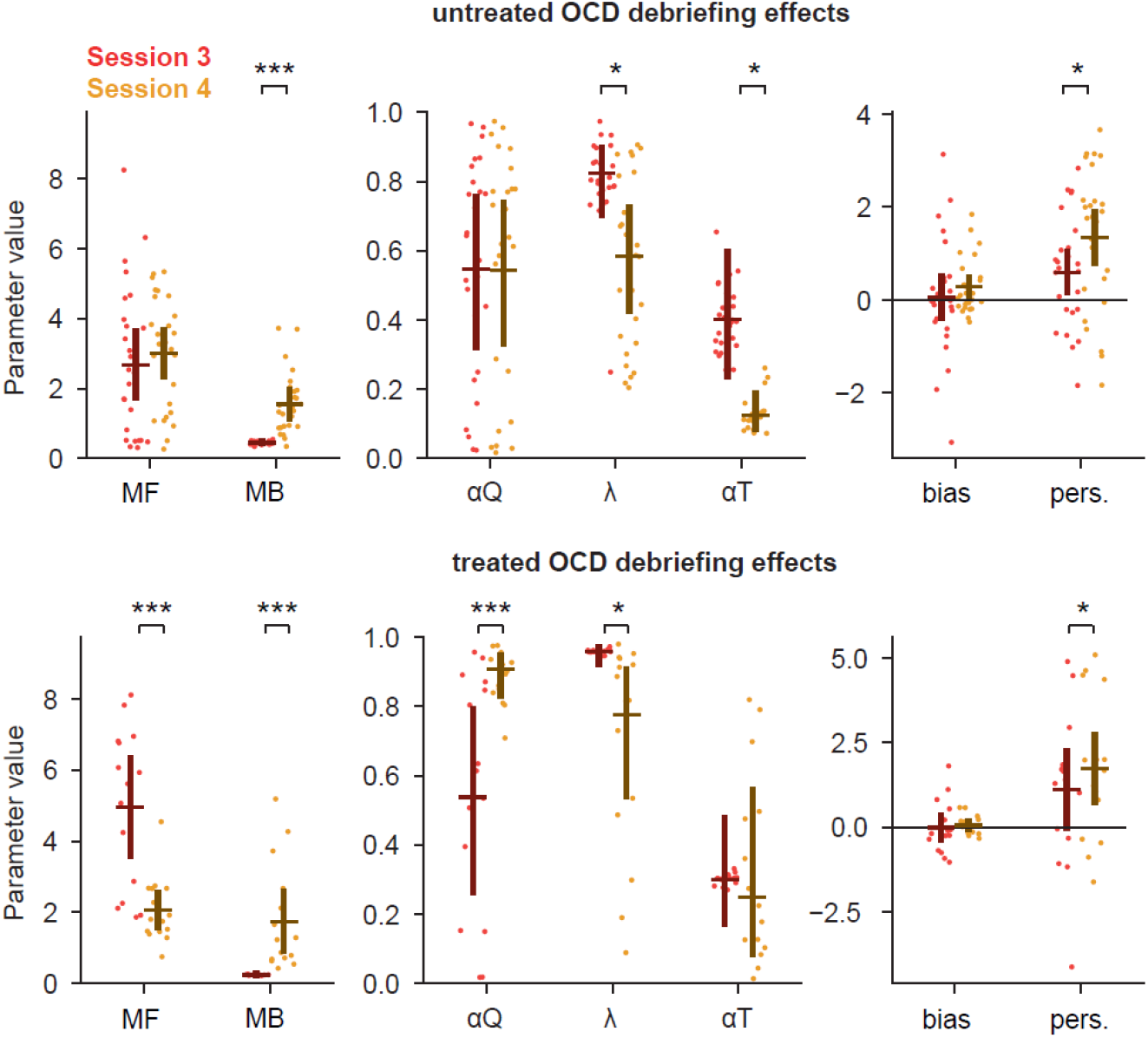
Comparison of debriefing effects in individuals with OCD between Lisbon and New York Samples. For each RL model parameter, dots indicate maximum a posteriori model parameter loading for individual subjects, bars indicate the population mean and 95% confidence interval on the mean. For each model parameter, permutation tests were used to assess effect of debriefing on the fitted loadings, separately for individuals with OCD recruited in New York (n=30), who were tested in the absence of pharmacological treatment (top panel), and in Lisbon (n=16), the majority of whom were under pharmacological treatment (bottom panel). These analyses were not performed in the remaining groups, because significant group-debriefing interactions were found only for the OCD sample (see Supplementary table sT1-3). Regarding the two variables for which such interactions were significant (*P*=0.04 for both; Supplementary table sT2), debriefing was found to reduce the strength of model-free RL (MF) and increase the value learning rate (αQ) in treated, but not untreated, individuals with OCD. RL model parameters: MF, Model-free strength; MB, Model-based strength; αQ, Value learning rate; λ, Eligibility trace; αT, Transition probability learning rate; bias, Choice bias; pers., Choice perseveration.

**Supplementary figure S5.**
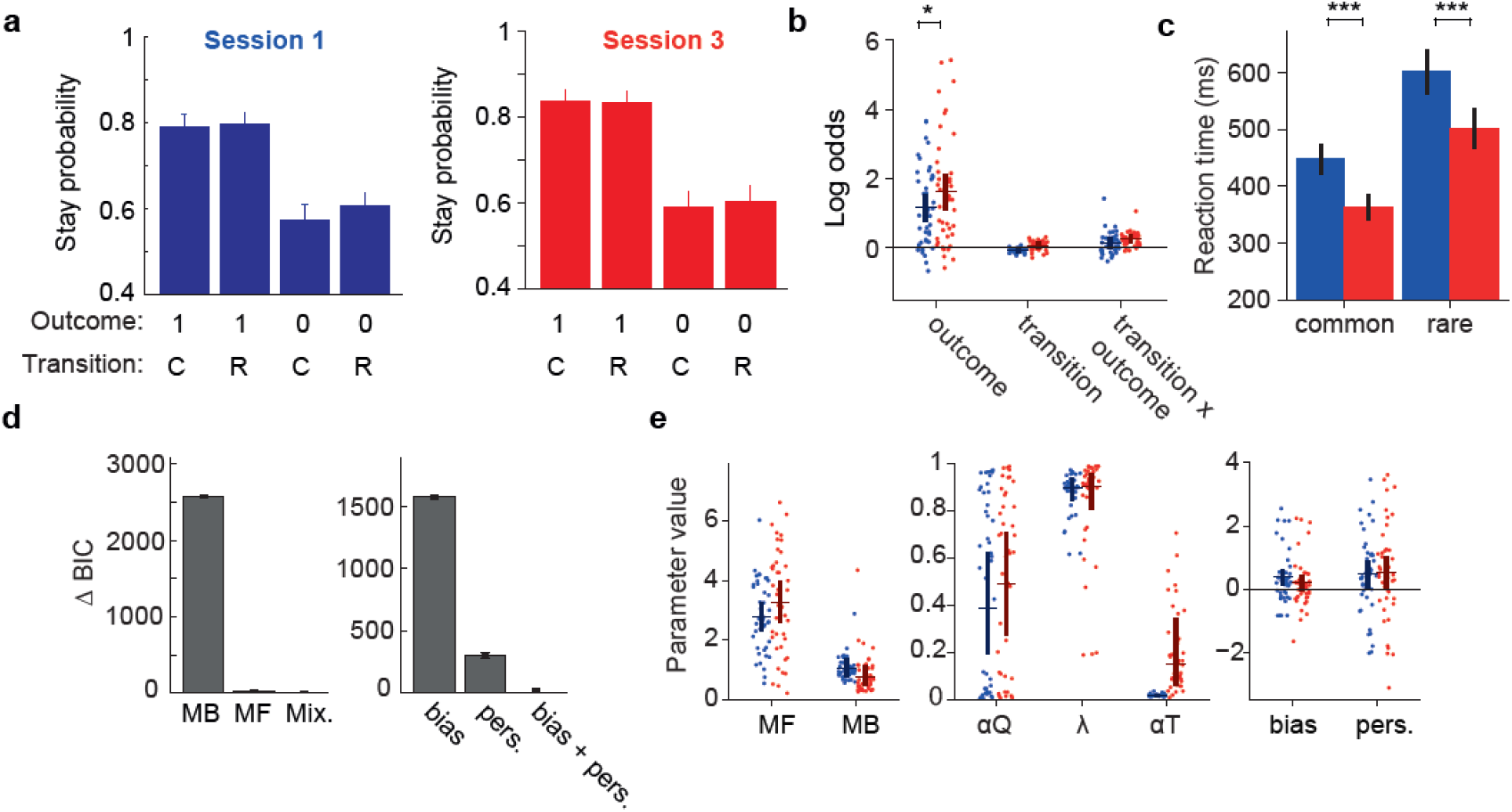
Learning effects in the Changing transition probabilities task. **a)** Stay probability analysis showing the probability of repeating the first step choice on the next trial as a function of trial outcome (rewarded or not rewarded) and state transition (common or rare). Error bars indicate the cross subject standard error (SEM). The left panel shows data from the first session, the right panel shows data from session 3. **b)** Logistic regression analysis of how the outcome (rewarded or not), transition (common or rare) and their interaction, predict the probability of repeating the same choice on the subsequent trial. Positive loading on the ‘outcome’ predictor indicates a tendency to repeat rewarded choices. Positive loading on the ‘transition’ predictor reflects a tendency to repeat choices followed by common transitions. Positive loading on the ‘transition x outcome’ interaction predictor indicates a tendency to repeat choices that were rewarded following a common transition, or that were not rewarded following a rare transition. Dots indicate maximum a posteriori loadings for individual subjects, bars indicate the population mean and 95% confidence interval on the mean. Statistical significance of differences in factor loadings for each predictor between session 1 (blue) and 3 (red) were evaluated using permutation tests. **c)** Mean first-step choice trajectories around reversals. In this and all panels, blue indicates session 1 while red indicates session 3. Dashed lines show exponential curves fitted to the average trajectories to obtain estimates of the time-course of learning following reversals. Confidence regions (mean ± cross subject standard error) are represented by shaded areas. **d)** Bayesian Information Criteria (BIC) model comparison for sessions 1-3. Left panel, comparison of model-based (MB), model-free (MF) and mixture (MF+MB) models. Right panel, comparison of mixture model with bias parameter, perseveration parameter, and bias + perseveration parameters. **e)** Comparison of mixture model fits between session 1 and session 3. RL model parameters: MF, Model-free strength; MB, Model-based strength; αQ, Value learning rate; λ, Eligibility trace; αT, Transition probability learning rate; bias, Choice bias; pers., Choice perseveration.

**Supplementary figure S6.**
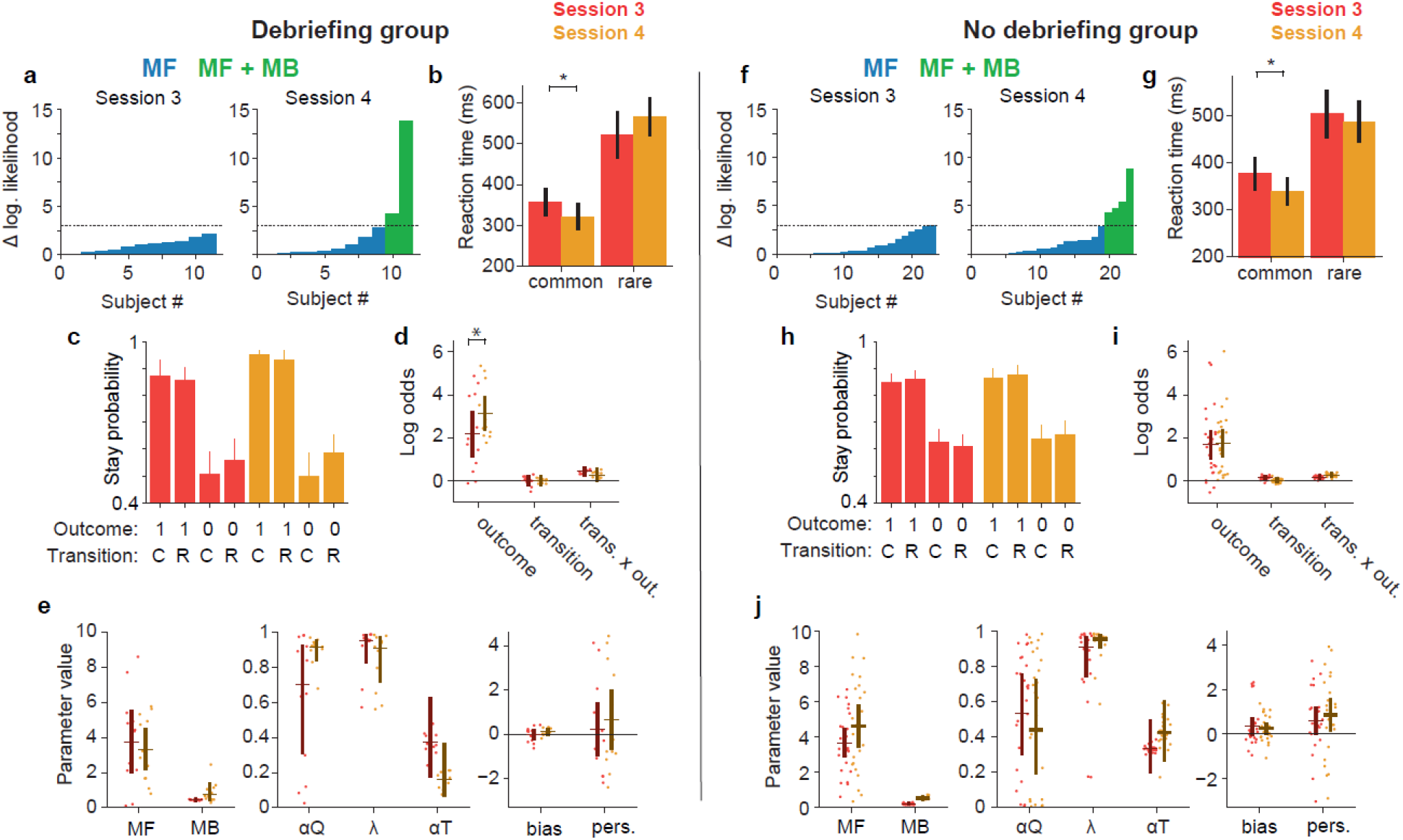
Effects of explicit knowledge in the Changing transition probabilities task. **(a, f)** Per-subject likelihood ratio test for use of model-based strategy on session 3 (left panel) and session 4 (right panel). Data was analysed separately for groups with (A) and without (F) debriefing. Y-axis shows difference in log likelihood between mixture (model-free + model-based) RL model and model-free only RL model. Blue bars indicate subjects for which likelihood ratio test favours model-free only model, green bars indicate subjects for which test favours mixture model, using a p<0.05 threshold for rejecting the simpler model. We compared sessions 3 and 4 only in the subjects for whom a likelihood ratio test indicated that model-based RL was not used in session 3. **(b, g)** Mean first-step choice trajectories around reversals. In these and all remaining panels, red indicates session 3 (before instruction) while gold indicates session 4 (after instruction). Dashed lines show exponential curves fitted to the average trajectories to obtain estimates of the adaptation time-course of learning following reversals. Confidence regions (mean ± across subject standard error) are represented by shaded areas. **(c, h)** Stay probability analysis showing the probability of repeating the first step choice on the next trial as a function of trial outcome (rewarded or not rewarded) and state transition (common or rare). Error bars indicated the cross subject standard error of the mean (SEM). In each group data was analysed separately for session 3 (red graph) and session 4 (gold graph). **(d, i)** Logistic regression analysis of how the outcome (rewarded or not), transition (common or rare) and their interaction, predict the probability of repeating the same choice on the subsequent trial. **(e, j)** Comparison of mixture model fits between session 3 (red) and session 4 (gold) in the group without instruction (left panels) and the group with instruction (right panels). RL model parameters: MF, Model-free strength; MB, Model-based strength; αQ, Value learning rate; λ, Eligibility trace; αT, Transition probability learning rate; bias, Choice bias; pers., Choice perseveration.

## Information provided to study participants

### A) Information before task

“You will now play a game in order to gain of as many rewards as possible.

Rewards will be represented in the screen as coins. Every time you get a coin, it will show up in the screen and it will be added to your total number of rewards. The number of coins you get will determine the value of the gift-card that you will receive at the end of your participation.

You will perform 1200 trials and in each trial you can get either one coin or no coin. At the end of those 1200 trials, 400 will be randomly chosen to count the final number of coins.

The minimum amount of money in your gift card will be 10 euros. For each coin that you get above 150 coins, you will get an increase of 20 cents in your gift card. Therefore, if you get 175 coins the amount will be 15 euros, 200 coins correspond to 20 euros and 225 coins correspond to the maximum amount that the gift-card can have, which is 25 euros. Amounts will be distributed rounded to the closer multiple of 5 euros.

At the top left corner of the screen, there will be a coin counter which shows how many coins you got in each session. That number may not have direct correspondence with the final amount, since that amount will be calculated using a random sample of trials.

You will play the game using the arrow keys after stimuli show up in the screen.

Each session of the game will last for approximately 15 minutes. Once the session is completed, a sentence thanking you for your participation will show up in the screen. When that screen shows up you should leave the room.

### B) Debriefing – Fixed transition probabilities version

We will now explain the structure of the game.

First the two central circles (upper and lower) are yellow, indicating that you can choose one of them.

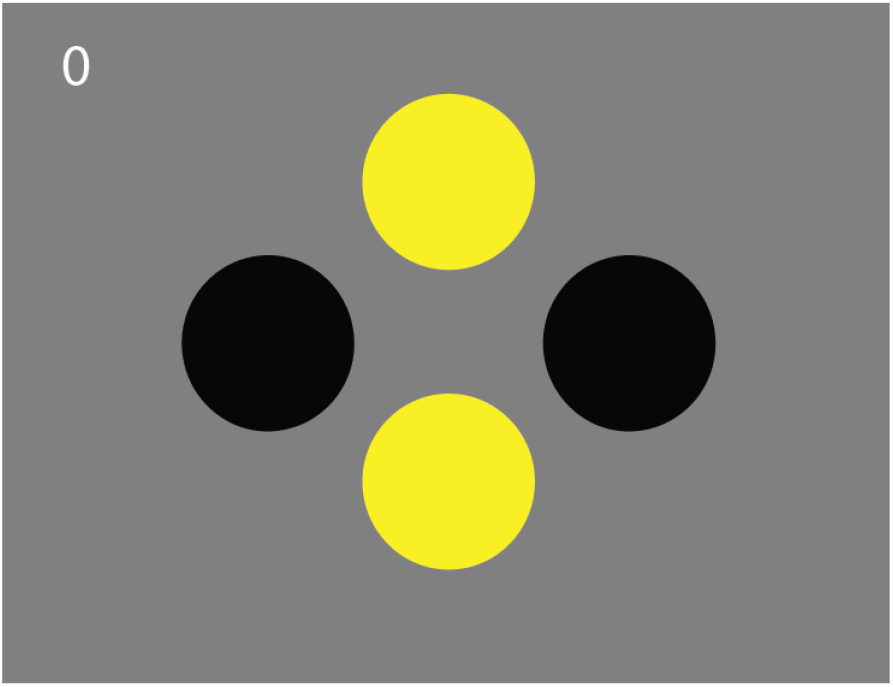

If you press the upper arrow key, you will choose the upper circle. If you press the lower arrow key, you will choose the lower circle.

After you choose the upper or the lower circle, one of the two side circles will light up, i. e., will turn yellow (left or right). After you press the arrow key that corresponds to the lateral circle that lit up (left or right), a coin may or may not appear.

The probability according to which the central circles give access to either one of the lateral circle also follows some rules.

If you choose the upper circle, one of two different things can happen. Most of the times (actually 80% of the times) the right side circle will light up. Rarely, the left side circle will light up.

If you choose the lower circle, most of the times (actually 80% of the times) the left side circle will light up. On the remaining occasions, the right side circle will light up.

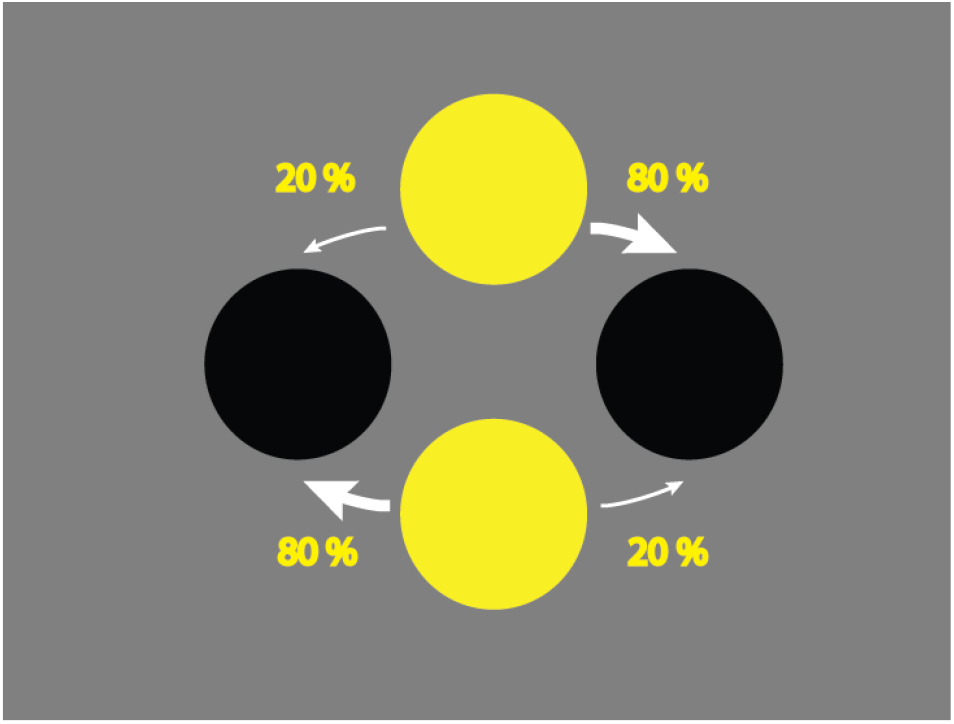

The left and right circles give access to the rewards, which are symbolized as coins. However, the probability of winning a coin is not equal on the left or on the right: it is always higher on one of the sides.

Sometimes it is higher on the left and sometimes it is higher on the right. The side in which that probability is higher changes after 20 or more trials.

You will now play a last session, with the same rules. Good luck!

### C) Debriefing – Changing transition probabilities version

We will now explain the structure of the game.

First the two central circles (upper and lower) are yellow, indicating that you can choose one of them.

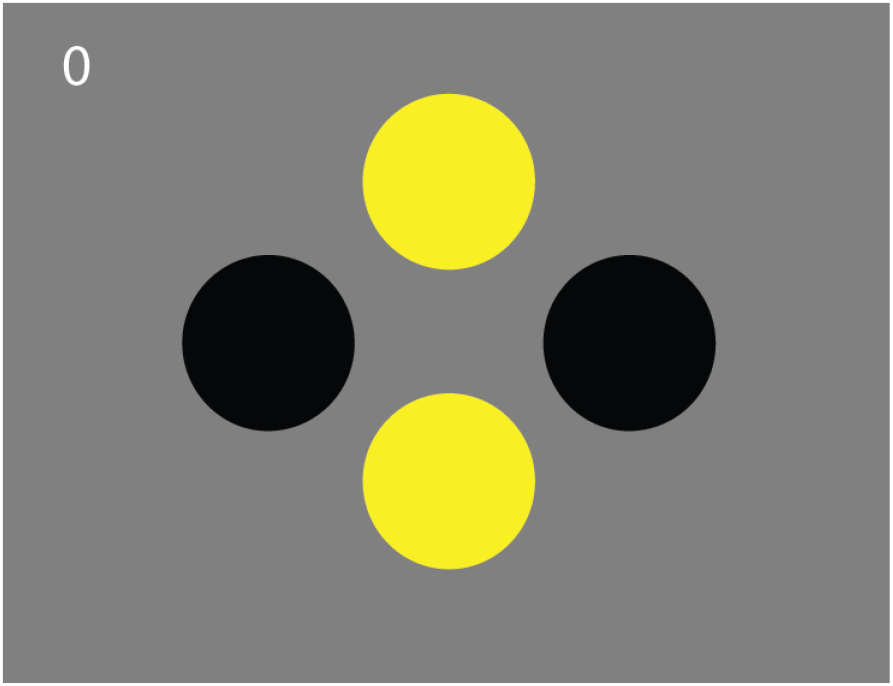

If you press the upper arrow key, you will choose the upper circle. If you press the lower arrow key, you will choose the lower circle.

After you choose the upper or the lower circle, one of the two side circles will light up, i. e., will turn yellow (left or right). After you press the arrow key that corresponds to the lateral circle that lit up (left or right), a coin may or may not appear.

The probability according to which the central circles give access to either one of the lateral circle also follows some rules. The game is divided in two types of blocks.

In “A” blocks, choosing the upper circle leads more frequently (80% of the times) to the lighting up of the right side circle. On the other hand, in these blocks, choosing the lower circle, leads more frequently (80% of the times) to the lighting up of the left side circle.

In “B” blocks, choosing the upper circle leads more frequently (80% of the times) to the lighting up of the left side circle. On the other hand, in these blocks, choosing the lower circle, leads more frequently (80% of the times) to the lighting up of the right side circle.

Therefore, in “A” blocks, if you choose the upper circle, one of two things can happen. Most of the times (actually 80% of the times), the right side circle will light up. Rarely (20% of the time), the left side circle will light up.

In these same “A” blocks, if you choose the lower circle, one of two things can happen. Most of the times (actually 80% of the times), the left side circle will light up. Rarely (20% of the time), the right side circle will light up.

Schematic representation of the structure of “A” blocks:

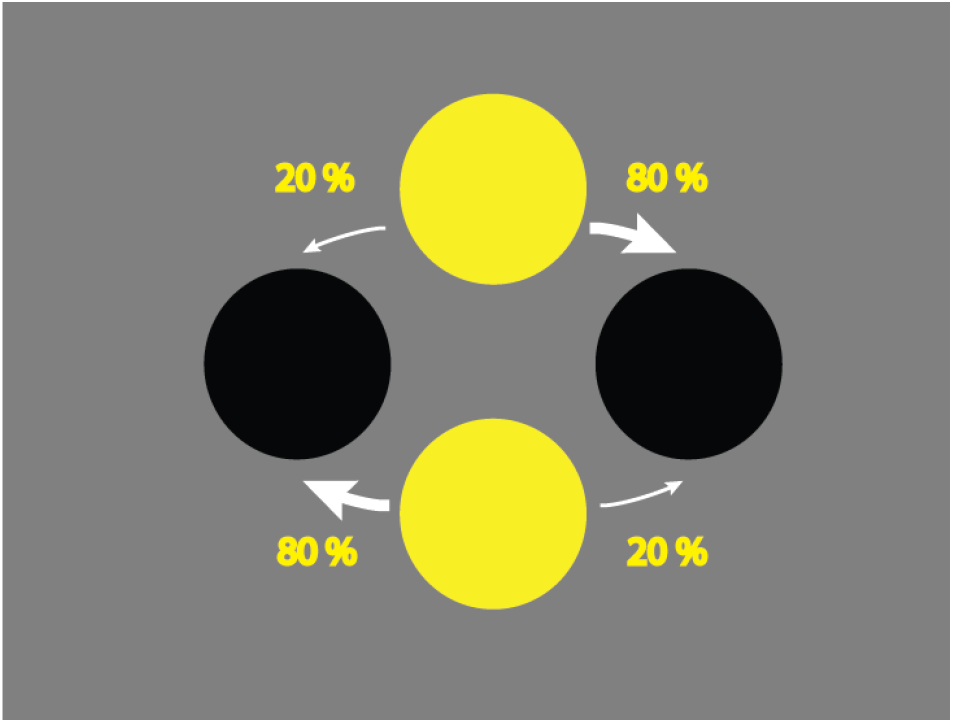

In “B” blocks, if you choose the upper circle, one of two things can happen. Most of the times (actually 80% of the times), the left side circle will light up. Rarely (20% of the time), the right side circle will light up.

In these same “B” blocks, if you choose the lower circle, one of two things can happen. Most of the times (actually 80% of the times), the right side circle will light up. Rarely (20% of the time), the left side circle will light up.

Schematic representation of the structure of “B” blocks:

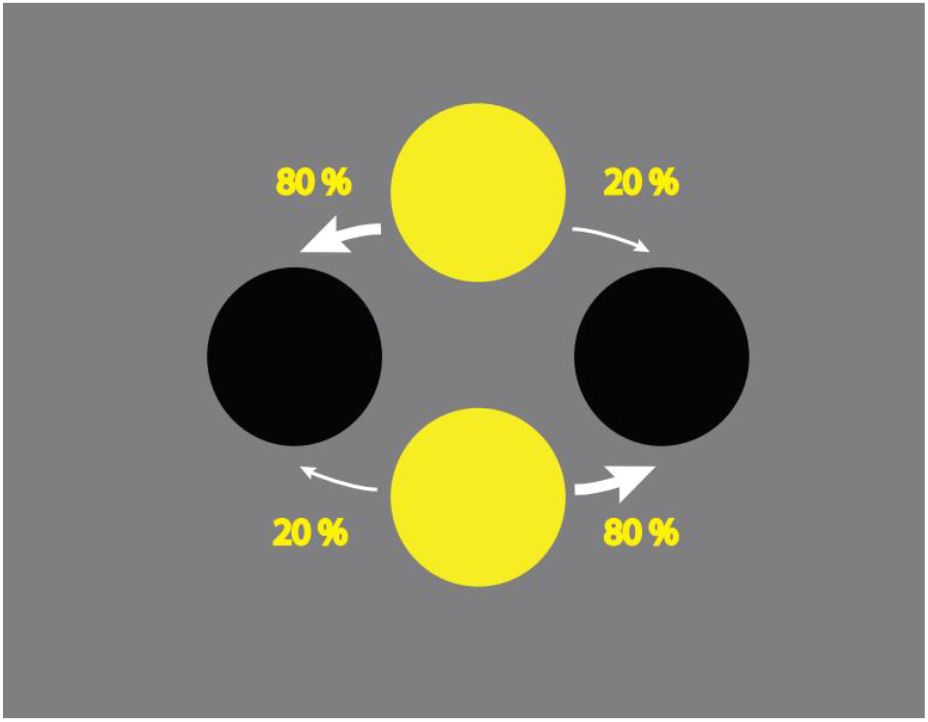

“A” blocks and “B” blocks alternate between them after 20 or more trials.

The left and right circles give access to the rewards, which are symbolized as coins. However, the probability of winning a coin is not the equal on the left or on the right: it is always higher on one of the sides. Sometimes it is higher on the left and sometimes it is higher on the right. The side in which that probability is higher changes after 20 or more trials.

You will now play a last session, with the same rules. Good luck!

